# Mind the Gaps: Validating species presence under sparse genomic recovery in clinical metagenomics

**DOI:** 10.64898/2026.09.23.26363603

**Authors:** Mahboobeh Behruznia, Frederick Silva-Toro, Diego A. Márquez

**Affiliations:** Department of Microbes, Infection and Microbiomes, School of Infection, Inflammation and immunology, College of Medicine and Health, University of Birmingham, Edgbaston, Birmingham B15 2TT, UK; Department of Mathematics, Faculty of Sciences, University of Chile, Ñuñoa, Santiago 7750000, Chile; School of Biosciences, University of Birmingham, Edgbaston, Birmingham B15 2TT, UK

**Keywords:** pathogen detection, long-read sequencing, taxonomic validation, low-biomass samples, microbial diagnostics

## Abstract

Clinical metagenomic sequencing is increasingly used to detect pathogens directly from clinical samples, but sparse and uneven genome recovery caused by low microbial biomass and stochastic fragment loss makes species-level validation difficult. Under these conditions, the same observed coverage may arise either from genuine genome-wide representation or from reads confined to conserved loci shared among related taxa. Current heuristic thresholds based on read counts, breadth or related alignment summaries target the realised coverage rather than the generating process. We show that, under hidden fragment loss, coverage is the wrong inferential object for uniformly valid species-level inference. What remains informative is the spatial dispersion of surviving reads across genomic coordinates. This change of inferential object allows us to ask how much of the candidate genome must be represented in any source capable of reproducing the observed evidence. This yields the Metagenomic Alignment Validation Index (MAVI), a conservative lower bound on the fraction of the candidate genome required to explain the observed read dispersion. MAVI therefore provides an error-controlled, interpretable measure of genomic confidence: the minimum genomic content a competing source would need to share with the candidate to mimic the observed evidence. Evaluated across 643 long-read metagenomic samples, including 280 clinical samples, MAVI reproduced independently supported organism calls, resolved sparse low-coverage ambiguities and exposed workflow-level collapse of stochastic dispersion, without relying on heuristic coverage thresholds.

## 2 Main

Clinical metagenomics is emerging as a powerful approach for identifying pathogens directly from clinical samples, as conventional diagnostic methods are often slow, presumptive, or inconclusive. However, sequencing datasets often display highly uneven and sparse coverage due to variation in pathogen load, predominant host derived DNA and substantial stochastic losses occur during host depletion, nucleic acid extraction and library preparation^1–4^. These factors make species level validation a central challenge in clinical metagenomics^4^, as limited and variable coverage complicates the distinction between genuine genome-wide presence and incidental matches within conserved regions^5^. These constraints are inherent to many clinical sample types, regardless of the tissue or fluid of origin, reflecting practical limits on biomass and recoverable genomic material in real diagnostic workflows^1^. In practice, this makes metagenomic validation an inverse problem: we observe a stochastically truncated genomic signal and must infer whether the full genome was physically present.

The identification of microbial species from long-reads metagenomic data typically relies on a multi-step process^4,6^ involving sample preparation, taxonomic assignment, sequence read alignment, and a final stage of validation (Figure 1). Fast k-mer–based classifiers, such as Kraken2^[7]^/KrakenUniq^8^, Centrifuge^9^ and CLARK^10^, play an essential role in detecting candidate taxa by rapidly comparing sequencing reads against reference databases. However, while these methods are efficient for generating a list of candidate taxa, they are not designed to perform species-level validation. In particular, they do not distinguish between true genome-wide presence and spurious matches arising from shared or conserved regions and therefore require an explicit downstream validation stage.

**Figure 1.**
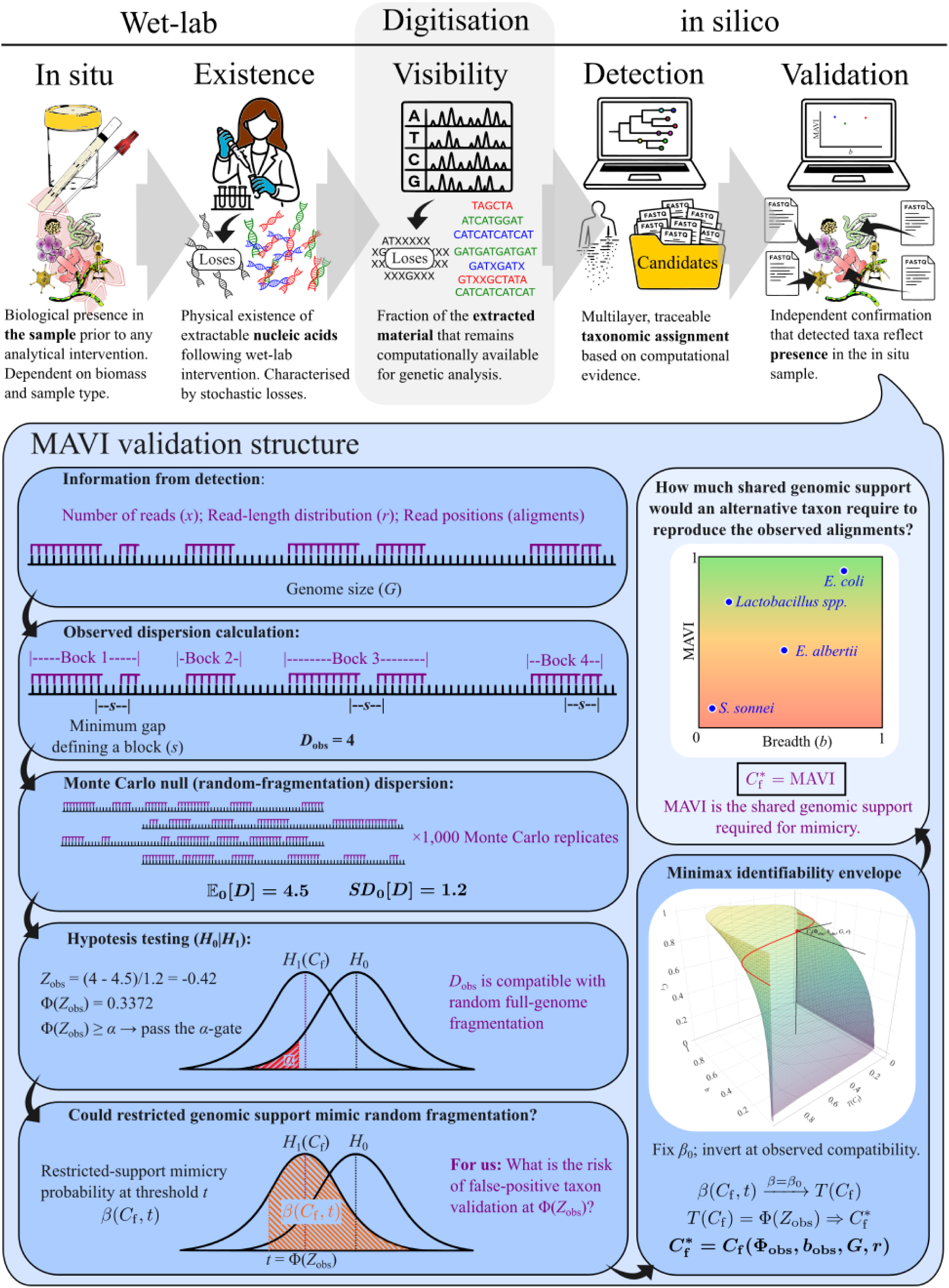
| Conceptual overview of the long-read metagenomic workflow and the Metagenomic Alignment Validation Index (MAVI). The upper panel summarises the progression from biological presence in the in situ sample through wet-lab extraction, digitisation and computational taxonomic detection. Each step can reduce the biological material that remains observable, so taxonomic detection produces candidate taxa that still require independent validation against the original sample. The lower panel shows the MAVI validation structure applied to each candidate. Observed read count, read-length distribution and genomic positions are used to calculate spatial dispersion, which is compared with a candidate-specific Monte Carlo reference representing random fragmentation across the full genome. The resulting compatibility coordinate, Φ(*Z*_obs_), can be expressed as a binary *α*-gate for full-support compatibility, while MAVI retains the continuous compatibility information. For a candidate support fraction *C*_f_, *β*(*C*_f_,*t*) quantifies restricted-support mimicry at threshold *t*. Fixing the tolerated mimicry probability *β*_0_ defines the minimax compatibility boundary *T*(*C*_f_); inversion of this boundary at the observed compatibility yields MAVI: the minimum shared genomic support required for an alternative taxon to mimic the observed dispersion compatibility.

Alignment-based methods, most commonly implemented with tools such as minimap2^[11]^ for long-read data, provide the subsequent validation stage applied to the candidate taxa identified by classifiers. By mapping reads back to reference genomes, these methods provide a more direct line of evidence for assessing candidate species. The resulting alignments enable calculation of metrics such as breadth of coverage, depth, and alignment identity. In an ideal scenario, validation is straightforward: full mapping across the entire reference genome provides unambiguous confirmation of presence, and the detection of species-specific unique genes during the alignment process also offers strong supporting evidence.

In practice, however, metagenomic datasets in clinical samples rarely achieve the levels of coverage required for full genome reconstruction. Breadth values below 50% are common, and complete recovery is exceptionally uncommon outside of dominant taxa^4^. Under such conditions, signals such as the detection of unique genes become difficult to interpret: their presence is informative, but their absence is inconclusive because limited coverage substantially reduces the probability of observing them at all^12,13^.

False-negative results are a recurrent challenge in clinical metagenomics, particularly when sequencing depth is low or host DNA dominates the sample. Even when a pathogen is physically present and detected as a taxonomic candidate, low and uneven genome coverage may provide insufficient evidence to validate that the observed match reflects genuine species-level presence^2^. Large multicentre evaluations have shown that most false negatives arise from stochastic losses during extraction and library preparation rather than from bioinformatic misclassification^1,5^. These losses reflect unavoidable physical and biochemical constraints inherent to current clinical sampling and processing workflows rather than procedural shortcomings.

These limitations are well recognised^4,14^, and a range of practical workarounds has emerged to manage them. Most commonly, studies apply minimum thresholds to decide whether a species should be considered present, including criteria based on relative read abundance, breadth of coverage, numbers of mapped reads, or cut-offs for alignment depth and identity. Such criteria can provide internal consistency within a given workflow, but vary widely across studies and often behave unreliably when coverage is low^5^.

A similar limitation affects methods that interrogate other features of the same incomplete evidence. YACHT, for example, formulates genome detection as a hypothesis test on sequence similarity, using ANI-based expectations to evaluate whether observed k-mer matches are consistent with a given reference^15^. A small number of reads originating from conserved loci may provide sufficient matching k-mers to satisfy an ANI-based test despite offering no evidence of genome-wide representation, whereas the loss of those fragments may yield a false negative even when the organism is present. More exhaustive strategies, including gene-level inspection, extensive cross-comparisons and large-scale computational filtering, can increase confidence but remain constrained by feasibility^5^. They nevertheless interrogate the same incomplete and uneven evidence without resolving the underlying uncertainty.

Other approaches, including that of Olm, et al. ^16^ and Sanguineti, et al. ^17^, used the spatial distribution of mapped reads as an additional feature beyond simple read counts in short-read metagenomics. By distinguishing broadly distributed alignments from locally concentrated matches, these approaches showed that read placement can improve taxonomic discrimination in Illumina datasets^12^. Their final decisions nevertheless relied on fixed or fitted thresholds, producing useful empirical classification rules rather than a general inferential framework.

Taken together, the field has relied on fixed, fitted or ad hoc thresholds as pragmatic validation criteria. The core difficulty lies in the simple fact that validation must be carried out using an incomplete and uneven representation of the genome. Any alignment-based decision is therefore only as informative as the genomic fragments that happen to survive sampling and become available for analysis. This creates a fundamental ambiguity: genuine genome-wide presence may appear weakly supported because large portions of the genome are missing, whereas a restricted set of conserved regions may produce apparently convincing support for a taxon that is not genuinely represented. Existing thresholds are consequently tied to the coverage and fragment-length characteristics of the workflows in which they were developed and do not provide a principled way to resolve this ambiguity. The problem is especially acute in clinical settings, where sparse recovery is inherent to the sample, uncertainty can affect treatment decisions, and simply relaxing confidence thresholds is not an acceptable solution.

The sparse and uneven genomic representation underlying this uncertainty arises directly from the attrition of information during wet-lab sample preparation^12^. Host depletion, nucleic acid extraction and library preparation follow deterministic protocols but interact in ways that are effectively stochastic at the scale of individual loci. Consequently, some genomic regions pass through intact, while others are only partially retained or lost entirely for reasons unrelated to genome structure or bioinformatic processing. We therefore propose that physical attrition does not merely remove information: by acting stochastically across genomic coordinates, it leaves a spatial imprint in the surviving alignments that retains information about the underlying genomic representation.

### 2.1 Theoretical framework: generator-level identifiability under stochastic truncation

In practical terms, stochastic truncation creates a simple ambiguity. Reads assigned to candidate species *A* may originate from fragments distributed across a genuinely present species *A* genome, or from a restricted set of loci shared with species *B* but originating from species *B*. If these two mechanisms produce the same observed alignment evidence, no statistic calculated solely from that evidence, including mean depth, read count, breadth or a tuned threshold, can determine which explanation is correct^18^. Such metrics describe the realised matches but do not uniquely identify the biological source that generated them. Species-level validation therefore cannot be resolved simply by selecting a different coverage metric or threshold.

Formally, this is a generator-level identifiability problem (Box 1). The wet-lab workflow acts as a stochastic truncation of an underlying genomic generator: fragments are retained or lost before sequencing, so the resulting alignments may identify only a compatibility class of possible genome-level sources rather than the unique source that produced them. Márquez and Silva-Toro ^18,19^ provide the formal framework for this problem, showing that under hidden support restriction the surviving inferential information is carried by stabilised spatial dispersion and can be inverted into a conservative support-fraction bound. Uniform validity over unresolved support geometries leads to a minimax construction. The framework provides the generator-level dispersion entropy diagnostic^19^, which summarises the retained stochastic dispersion structure, and the Generator Identifiability Envelope^18^, which provides a conservative lower bound on the support fraction required to explain the observed dispersion. Here, we instantiate this framework on genomic coordinates and construct the corresponding minimax dispersion envelope for metagenomic validation.

The mechanistic basis of the framework is the classical Lander and Waterman ^20^ model of stochastic genome fragmentation. This model provides the full-support reference process: when fragments arise from across the candidate genome and survive stochastically, their positions remain distributed over the available genomic coordinates. If the underlying source is instead confined to an unknown genomic fraction, the possible spatial arrangements of surviving alignments are correspondingly constrained. Under sparse recovery, however, these regimes need not be perfectly separable: a restricted-support generator may by chance mimic the dispersion expected from genome-wide fragmentation. The inferential information therefore lies not merely in whether the alignments appear dispersed, but in how much genomic support an alternative generator would require to reproduce that dispersion.

Accordingly, we separate metagenomic validation into two questions. First, are the mapped reads compatible with random fragmentation across the full candidate genome? Second, if they are, how much genomic support would a competing restricted source require to reproduce the observed dispersion?

We formulate this through the hypotheses

*H_0_: the observed reads arise through random fragmentation over the full candidate genome;*
*H_1_(c): the observed reads arise from an unknown restricted support of total length at most cG.*

where *G* is the candidate genome length. For each candidate support fraction *c*, and compatibility threshold *t*, we quantify the worst-case probability that any admissible restricted-support alternative attains compatibility above *t* with full-support fragmentation. This defines the restricted-support mimicry probability over the unresolved support geometries. Fixing the tolerated mimicry probability *β*_0_ converts this worst-case profile into a minimax compatibility boundary *T*(*c*). Inverting that boundary at the observed dispersion-compatibility coordinate yields the genomic specialisation of the *Generator Identifiability Envelope* (GIE), which we define here as the *Metagenomic Alignment Validation Index* (MAVI).

Operationally, MAVI is obtained from Monte Carlo fragmentation models^21,22^ parameterised by the candidate genome, observed read count and empirical read-length distribution. MAVI yields a continuous support-fraction bound with a direct mechanistic interpretation: the minimum fraction of the candidate genome that a restricted-support alternative must access to mimic the observed dispersion compatibility. Full statistical and mathematical details are provided in Supplementary Note 1.

#### Box 1

**– Formal non-identifiability of genomic source under incomplete observation**

Suppose that the observed alignment evidence consists of genomic regions *A*1, *A*2, *A*3, *A*4, and that the candidate genomes contain the following subsets:

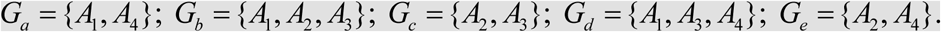

The observed regions can then be accounted for by several distinct source configurations. For example, *G_a_* ∪ *G_c_* = {*A*_1_, *A*_2_, *A*_3_, *A*_4_}, while *G_b_* ∪ *G_d_* = {*A*_1_, *A*_2_, *A*_3_, *A*_4_}. These configurations are biologically different, yet both remain compatible with the recovered evidence. The arises whenever different combinations of organisms, shared loci and fragment-survival patterns can explain the same incomplete observation.

More generally, let Γ denote a latent genomic source configuration and let χ(Γ) denote the observations that it can generate. For an observed alignment set *X*_obs_, the admissible compatibility class is

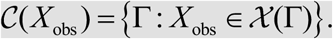

If

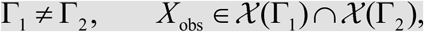

then the realised evidence does not uniquely identify which source configuration generated it. One explanation may nevertheless be selected using assumptions that are neither testable from nor supplied by the observed evidence. Such a selection does not remove the remaining compatible configurations.

The observed evidence may also be compatible with unmodelled organisms, mixtures of organisms, shared-locus configurations or other unconsidered sources. The analyst therefore cannot know whether the candidates examined exhaust the biologically possible explanations.

Once multiple source configurations remain compatible with the observation, the data do not identify which one generated it. A classical analysis can return a unique answer only by imposing assumptions that are not testable from the observed evidence. Those assumptions select an explanation; they do not make it identifiable from the data.

## 3 Mechanistic construction of the Metagenomic Alignment Validation Index

Our results show that genome-wide presence need not produce high coverage, but it does constrain the law of spatial dispersion of surviving alignments. Under full genomic support, stochastic fragmentation induces a candidate-specific dispersion law across genomic coordinates; restriction to shared loci alters that law by limiting the coordinates from which reads can arise. The practical consequence of this distinction is a sharp reduction from a large set of candidate taxa to a much smaller, well-defined set of validated ones. Before detailing the framework, we first show the distribution of MAVI across all candidate taxa in the clinical datasets and a representative sample-level analysis (Figure 2). MAVI inverts the observed compatibility with the full-support dispersion law to obtain a lower bound on the genomic fraction required for restricted-support mimicry. Because this bound is derived from the physical fragmentation-and-survival process rather than from heuristic coverage thresholds, it provides a mechanistically interpretable basis for validating metagenomic evidence.

**Figure 2.**
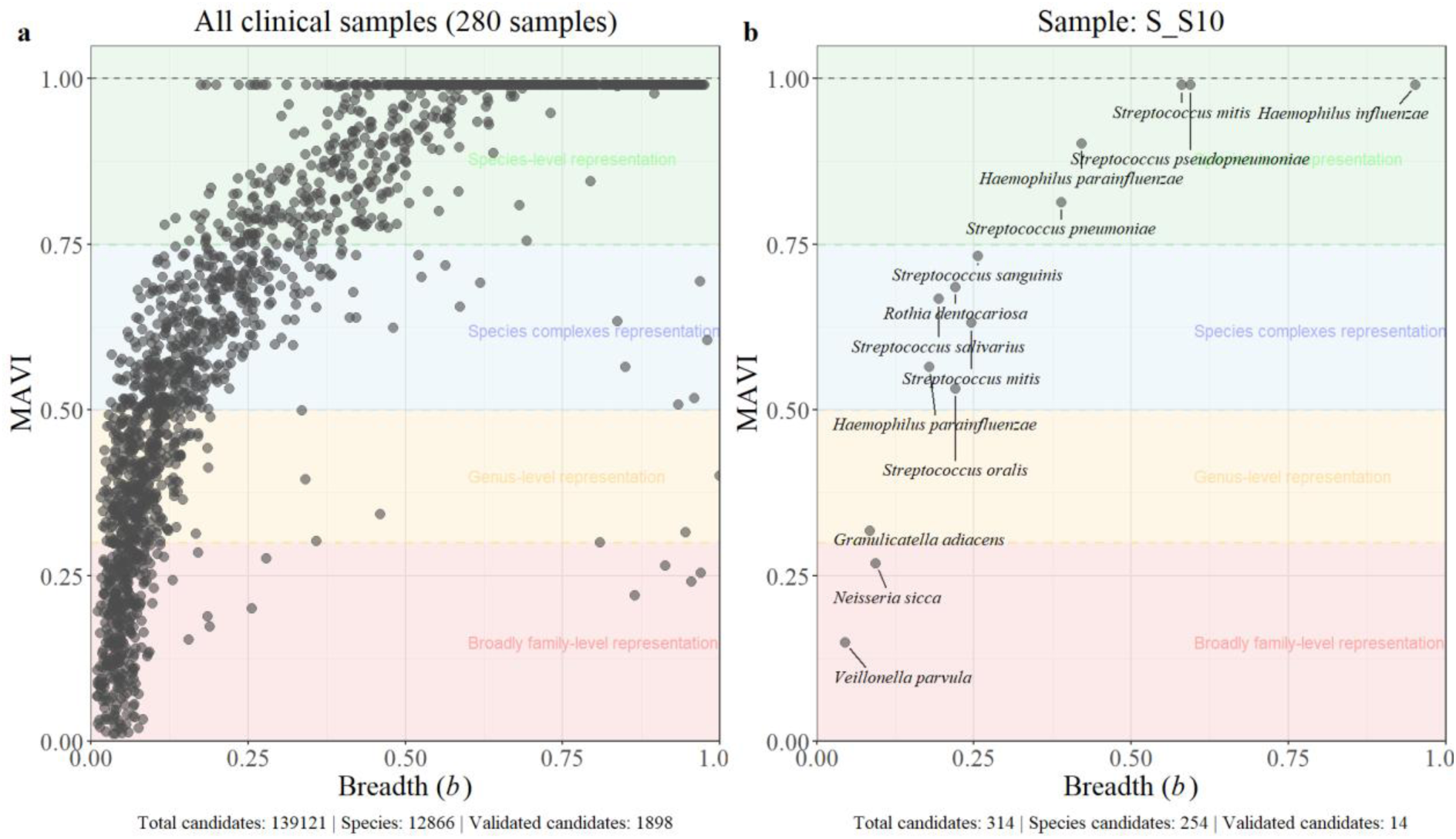
| Breadth–MAVI plots for all candidates and for a representative sample. (a) Breadth–MAVI distribution for all candidate genomes across the analysed datasets. Each point represents a candidate genome plotted by its observed breadth (*b*) and MAVI value. (b) Breadth–MAVI distribution for a representative sample (Charalampous, et al. ^23^ S_S10), with candidate genomes labelled by taxonomic assignment using Kraken2. Coloured background bands are provided solely as visual guides to facilitate reading of the plot; they do not represent hard taxonomic boundaries and are not used in the MAVI calculation or inference. Counts below each panel report the total taxonomic candidates from Kraken2, species-level candidates and MAVI-validated candidates.

MAVI values are interpreted directly on the genomic-support scale. A value of *m* means that any restricted-support explanation capable of mimicking the observed dispersion must access at least a fraction *m* of the candidate genome. High values therefore exclude explanations confined to small shared genomic regions, whereas low values leave such explanations viable and the species-level assignment unresolved. This distinction is most consequential precisely in the intermediate recovery regime, where classical coverage metrics remain non-identifying but the spatial dispersion of the surviving alignments can still constrain the minimum genomic support required by any competing explanation.

Figure 2 illustrates the discriminatory power of MAVI at both cohort and individual-sample scales. Across more than one hundred thousand candidate-genome alignments, MAVI reduced the plausible solution space to approximately two thousand statistically validated candidates, each assigned an explicit genomic support-fraction bound (Figure 2a). At the individual-sample level, hundreds of initial candidates were similarly resolved into a small set of well-supported genomes (Figure 2b). This reduction follows from the GIE principle^18^, mechanistically grounded in stochastic genome fragmentation, and is obtained under explicit control of both the compatibility gate and restricted-support mimicry (*α* = 0.001, *β*_0_ = 0.001).

MAVI operates on information already produced by taxonomic classification and alignment, adding no further alignment or database-search requirements. In our workflow, taxonomic candidates were identified using Kraken2, after which reads assigned to each candidate were aligned to the corresponding reference genome using minimap2. For each candidate, MAVI uses the genome size *G*, observed mapped-read count *x*, empirical read-length distribution *r*, and genomic coordinates of the alignments to perform candidate-specific Monte Carlo dispersion analysis. Genome breadth *b* is retained as an interpretable display coordinate but is not an independent source of inferential evidence. MAVI is therefore applied as a computationally lightweight post hoc validation step to all candidate taxa, without requiring prior filtering, reassembly or heuristic thresholding.

### 3.1 Stochastic read dispersion analysis

For each candidate organism, we quantify the spatial arrangement of aligned reads using the stabilised dispersion functional *D* ^[19]^, defined as the number of non-overlapping alignment blocks separated by more than the stabilisation scale *s*. Alignments whose genomic coordinates overlap or are separated by at most *s* = 1,000 bp are merged into a single block, whereas alignments separated by more than 1,000 bp contribute an additional block. This construction ensures that *D_s_* captures genuinely distinct genomic loci rather than local fluctuations caused by alignment jitter, short gaps or indels. As shown in Supplementary Note 2, *s* = 1,000 bp lies within the informative stabilisation regime^18^ for long-read data, and the behaviour of *D_s_* remains stable across a broad range of nearby values.

Biologically, *D_s_* records the number of spatially distinct genomic regions represented after local alignment structure has been collapsed. It therefore characterises the geometry of the evidence rather than its amount: two candidates with the same number of mapped reads may have very different *D_s_* values depending on whether those reads occupy one localised region or many separated loci. For a given candidate, applying *D*_s_ to the observed alignments yields the observed dispersion *D*_obs_. Because the attainable dispersion also depends on genome size, read count and read-length distribution, *D*_obs_ is not interpreted in isolation but relative to its candidate-specific random-fragmentation law. This converts the observed arrangement from a descriptive spatial pattern into a testable statement about the genomic support required to generate it. The use of positional dispersion is related to classical analyses of genomic randomness, including Karlin and Brendel ^24^, but here it is used to infer hidden genomic support under partial recovery.

To calibrate *D*_obs_, we instantiate the Lander-Waterman Poisson fragmentation process^20^ using candidate-specific inputs *x*, *r* and *G*. In each Monte Carlo iteration, *x* reads are placed uniformly across the candidate genome, with lengths sampled from the empirical distribution *r*, and the simulated alignments are collapsed using the same stabilisation rule used to calculate *D*_obs_. Repeating this procedure 1,000 times yields the candidate-specific full-support null distribution of *D_s_*. Its mean, _0_[*D_s_*], and standard deviation, SD_0_[*D_s_*], provide the reference against which *D*_obs_ is evaluated.

#### 3.1.1 Full-support compatibility: *Z*-score evaluation and the *α*-gate

The candidate-specific null distribution places *D*_obs_ on a common compatibility scale. We therefore standardise the observed dispersion as

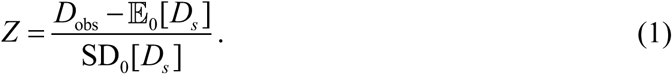

The resulting *Z*-score gives the signed position of the observed dispersion within its full-support random-fragmentation null. Values near zero indicate agreement with the null expectation, whereas increasingly negative values indicate less spatial dispersion than expected. Because each observation is standardised against the null conditioned on its own *G*, *x* and *r*, the resulting values are comparable across candidate genomes and recovery configurations.

We map the standardised dispersion to the continuous lower-tail coordinate Φ(*Z*). Because the relevant departure is reduced dispersion, a candidate fails the compatibility gate when Φ(*Z*) < *α* and passes when Φ(*Z*) ≥ *α*, with *α* = 0.001 (Figure 1). Here, Φ(*Z*) is derived from the candidate-specific Monte Carlo null and provides the common coordinate used for subsequent minimax inversion. Figure 3 summarises these values across all candidate taxa in the clinical datasets and illustrates their interpretation within an individual sample.

**Figure 3.**
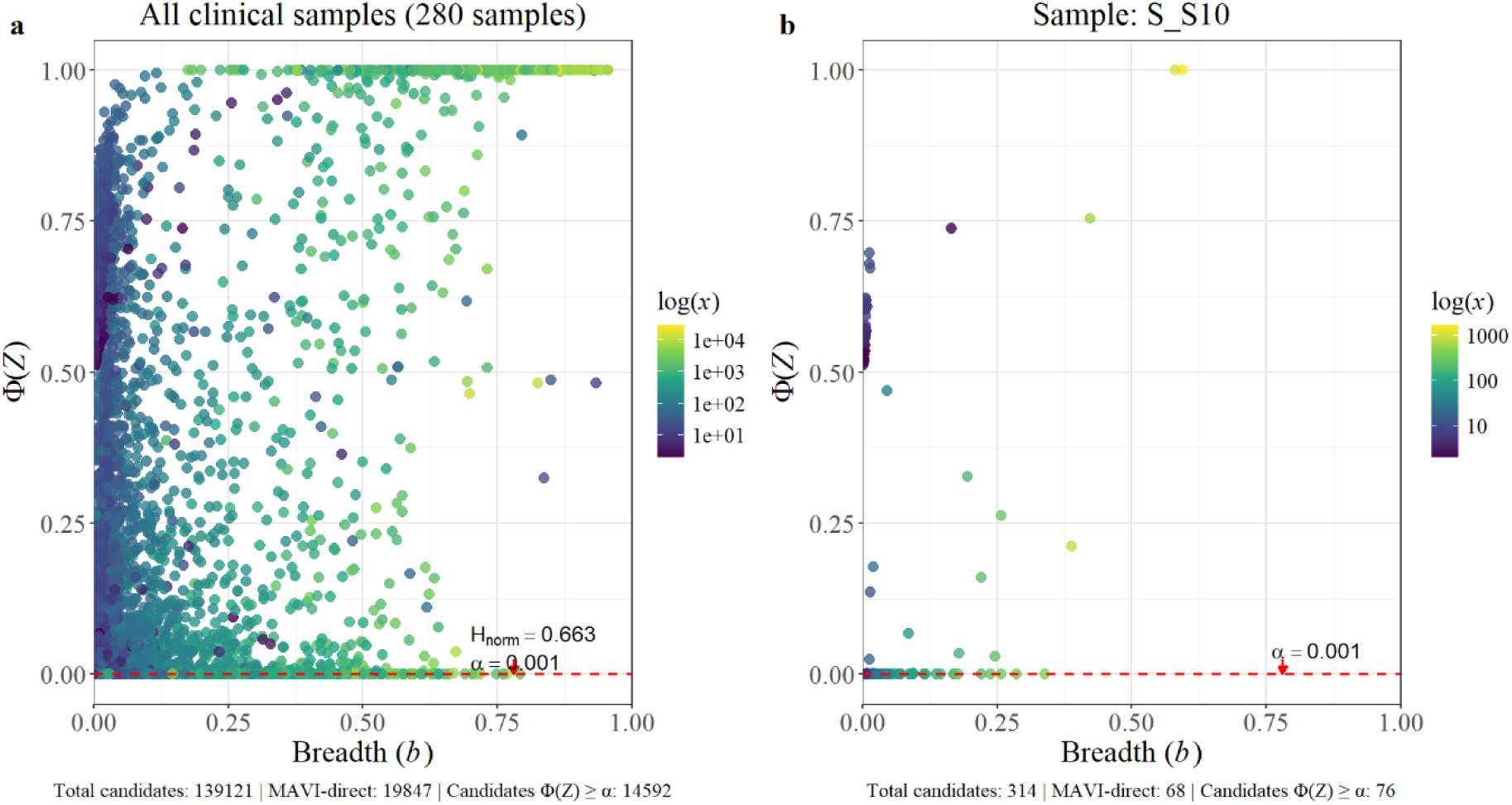
| Dispersion compatibility across all clinical samples and within a representative sample. The left panel shows all candidate genomes identified across 280 clinical metagenomic samples, whereas the right panel shows the candidate-level results for sample S_S10 from Charalampous, et al. ^23^. Each point represents a candidate genome, positioned according to its observed breadth *b* and dispersion-compatibility coordinate Φ(*Z*). Points are coloured by the number of assigned reads *x*, shown on a logarithmic scale. The dashed red line marks the full-support compatibility gate at *α* = 0.001. *H*_norm_ is the normalised generator-level dispersion entropy, calculated from the distribution of candidates satisfying Φ(*Z*) > *α*. Counts below each panel report the total taxonomic candidates, MAVI-direct candidates and candidates passing the *α*-gate. MAVI-direct candidates are those exhibiting Monte Carlo null-variance collapse; these candidates are carried forward directly to MAVI analysis.

Figure 3 shows that neither breadth nor read count alone determines compatibility with genome-wide random fragmentation. Candidates with similar breadth and read abundance span a wide range of Φ(*Z*), and some candidates with extensive coverage and many mapped reads nevertheless fall below the *α*-gate. This is the empirical manifestation of the identifiability problem formalised in Box 1: apparently strong coverage summaries may still arise from spatially concentrated alignments and cannot, by themselves, distinguish restricted genomic support from genuinely dispersed genome-wide representation.

### 3.2 Restricted-support mimicry

The dispersion test first excludes candidates whose alignments are incompatible with full-support random fragmentation. Passing this *α*-gate, however, does not certify genome-wide presence: under sparse recovery, a restricted-support generator may still produce sufficient dispersion to remain compatible with full-support fragmentation. We therefore quantify, for each candidate support fraction *C*_f_ and compatibility threshold *t*, the worst-case probability that restricted genomic support reproduces full-support compatibility above *t*. In other words, it asks how readily another taxon sharing only part of the candidate genome could mimic the evidence for that candidate. This restricted-support mimicry probability is the quantity carried forward into the minimax MAVI envelope (Figure 1).

To characterise this behaviour systematically, we simulate restricted-support alternatives by confining read placement to a fraction *C*_f_ of the candidate genome. For each combination of *G*, *C*_f_ and *r*, and across a grid of read counts *x*, reads are placed uniformly within a contiguous region of length *C*_f_*G*, with lengths drawn from *r*. Each realisation yields an empirical breadth *b*, a stabilised dispersion value *D*_s_, and the corresponding compatibility coordinate Φ(*Z*), calculated against the candidate-specific full-support null defined in Section 3.1.

For example, when *C*_f_ = 0.30, all *x* reads are placed uniformly at random within a region spanning 30% of the candidate genome, with no placements permitted in the remaining 70%. The resulting dispersion is then evaluated against the full-support null, in which reads may arise across the entire genome of length *G*. The simulations therefore quantify how often a restricted-support generator can mimic compatibility with genome-wide random fragmentation.

Biologically, restricted-support alignments may arise from conserved loci, shared gene families or mobile genetic elements that are not specific to the candidate genome^12^. The cluster fraction *C*_f_ therefore represents the maximum fraction of the candidate genome over which such an alternative source could generate compatible alignments.

This construction generates 100,000 Φ(*Z*) values for each restricted-support simulation setting. Indexed by *G*, *r*, *C*_f_, and the resulting breadth *b*, these outputs form an empirical lookup table characterising how restricted-support alternatives behave when evaluated against the same full-support null. For fixed *G* and *r*, the distribution at each (*b*, *C*_f_) coordinate is summarised by its upper (1-*β*_0_)-quantile. This defines the compatibility boundary *T*(*b* | *C*_f_) in the (*b*, Φ(*Z*)) plane.

Because *T*(*b* | *C*_f_) is defined by the upper (1-*β*_0_)-quantile, a restricted-support alternative occupying fraction *C*_f_ exceeds this boundary with probability at most *β*_0_. Observed compatibility below the boundary therefore cannot exclude that alternative at the declared tolerance, whereas compatibility above it rules out *C*_f_-restricted mimicry at level *β*_0_ (Supplementary Note 3). Biologically, *T*(*b* | *C*_f_) specifies how much apparent compatibility with genome-wide fragmentation can still be generated by alignments confined to fraction *C*_f_. At low breadth, where mimicry is easier, the boundary becomes correspondingly more stringent: sparse evidence is not rejected merely for being sparse, but restricted-support explanations are excluded only when the observed compatibility exceeds what they can reproduce.

Crucially, this ambiguity is not confined to low breadth. Sparse recovery makes restricted-support mimicry difficult to exclude, while extensive shared genomic support can produce broad yet spatially constrained alignments even at high breadth. Breadth is therefore non-identifying across its full range. Restricted-support simulations instead quantify, at each breadth, how strongly limited genomic supports can mimic full-support compatibility. Their upper (1-*β*_0_)-quantiles define the minimax compatibility boundaries that determine which restricted-support fractions remain admissible for the observed evidence.

### 3.3 Minimax compatibility boundaries at tolerance *β*_0_

We operationalise the restricted-support mimicry framework from Section 3.2 by fixing the tolerated probability of restricted-support mimicry at *β*_0_ = 0.001, corresponding to a one-sided confidence level of 1-*β*_0_ = 99.9% (Supplementary Note 4). For each realised breadth *b* and candidate support fraction *C*_f_, the boundary *T*(*b* | *C*_f_) is defined by the 99.9th percentile of Φ(*Z*) under the restricted-support simulations.

Compatibility above *T*(*b* | *C*_f_) therefore excludes mimicry by a *C*_f_-restricted alternative at 99.9% confidence, whereas compatibility below the boundary leaves that alternative unresolved. These boundaries identify which restricted-support fractions remain admissible for the observed compatibility and provide stringent protection against false validation through restricted-support mimicry, a particularly important requirement in clinical metagenomics^1^.

The family of boundaries *T*(*b* | *C*_f_) follows a smooth, monotonic transition with realised breadth (Figure 4). At low breadth, the compatibility distributions generated by full-support and restricted-support fragmentation overlap substantially, so a high compatibility value is required to exclude mimicry. As recovery increases, the admissible mimicry region contracts and *T*(*b* | *C*_f_) declines towards zero: a generator confined to fraction *C*_f_ has progressively less freedom to reproduce the dispersion generated across the full candidate genome. The resulting transition is sigmoidal, reflecting the rapid gain in discriminatory information once recovery moves beyond the regime of strongest overlap.

**Figure 4.**
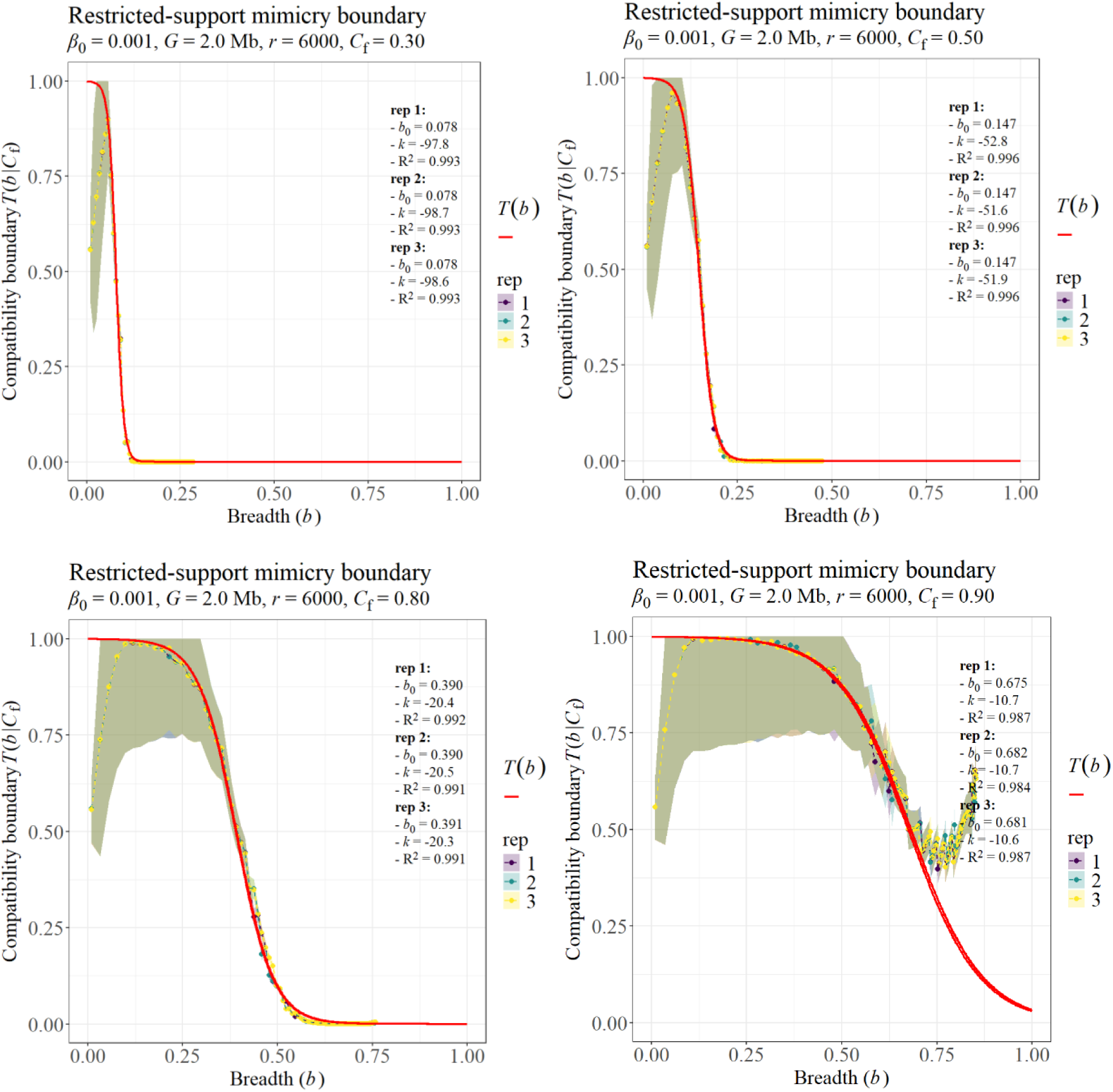
| Restricted-support mimicry boundaries and their analytical representation at *β*_0_ = 0.001. For each restricted-support fraction *C*_f_, alignments were simulated by placing reads within a contiguous genomic region of length *C*_f_*G* and evaluating their dispersion against the corresponding full-support reference process. At each realised breadth *b*, the empirical boundary *T*(*b*|*C*_f_) is given by the upper 1-*β*_0_ quantile of the restricted-support compatibility distribution. Observed compatibility above this boundary therefore excludes mimicry by a *C*_f_-restricted alternative at the declared tolerance *β*_0_ = 0.001. Panels show representative boundaries for *G* = 2 Mb, *r* = 6000 and *C*_f_ = 0.30, 0.50, 0.80 and 0.90, each evaluated in three independent simulation repetitions. Red curves show the logistic representation of *T*(*b*), with fitted midpoint *b*_0_, steepness *k* and deviance-based pseudo-R^2^ reported for each repetition. The near-complete overlap of the fitted curves shows that the boundary geometry is highly reproducible across independent simulations and can therefore be captured by a low-dimensional analytical representation. Shaded ribbons show the standard deviation of the underlying restricted-support compatibility values Φ(Z), rather than uncertainty in the fitted boundary. Each repetition comprised 10,000,000 simulated alignments, and each boundary point was estimated from 100,000 Monte Carlo realisations.

For each (*b*,*C*_f_), *T*(*b* | *C*_f_) specifies the compatibility level above which mimicry by a *C*_f_-restricted alternative is excluded at 99.9% confidence. Across more than 50 billion Monte Carlo realisations spanning broad ranges of genome size, read-length distribution and restricted-support fraction, these boundaries retained a highly reproducible sigmoidal geometry, with little variation in their fitted shape. This regularity reveals a stable, low-dimensional structure in the restricted-support mimicry surface. Crucially, it turns boundary estimation from a candidate-specific simulation problem into a modelling problem: rather than requiring millions of Monte Carlo iterations to reconstruct the boundary for every candidate, its behaviour can be learned from the simulation grid and subsequently evaluated through a lightweight analytical representation.

We therefore model *T*(*b* | *C*_f_) using a logistic form, replacing repeated simulation during candidate evaluation with a tractable analytical calculation. For fixed *G*, *r* and *C*_f_, we write this boundary as *T*(*b*):

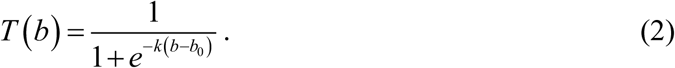

Here, *b*_0_ marks the midpoint of the compatibility transition and *k* determines its steepness. Together, they describe where restricted-support mimicry begins to lose viability and how rapidly that transition occurs as breadth increases. Technical details of the monotonic modelling interval are provided in Supplementary Notes 3.2–3.4.

As *C*_f_ increases, restricted-support generators gain access to a larger set of genomic coordinates and can reproduce a broader range of dispersion patterns. This behaviour is already visible at *C*_f_ = 0.90, where substantial mimicry remains possible even at high breadth: a competing source with access to 90% of the candidate genomic support leaves only the remaining 10% of genomic coordinates to distinguish the two explanations. The boundary *T*(*b* | *C*_f_) therefore rises and the admissible mimicry region expands. In the limit *C*_f_ → 1, the restricted-support process converges to the full-support null, and the two explanations become indistinguishable. This dependence on *C*_f_ shows why the evidential meaning of a given breadth cannot be universal.

At this stage, the two-stage error-control parameterisation becomes explicit: *α* controls the initial compatibility gate, whereas *β*_0_ controls restricted-support mimicry. We use *α* = 0.001 and *β*_0_ = 0.001, but the formulation supports any declared pair of error tolerances (Supplementary Note 4).

### 3.4 Metagenomic Alignment Validation Index (MAVI)

Across restricted-support fractions *C*_f_, the logistic boundaries defined in Eq. (2) form a smooth empirical surface in the (*C*_f_, *b*, *T*) space (Figure 5). For fixed *G* and *r*, each value of *C*_f_ defines one boundary *T*(*b* | *C*_f_); together, these boundaries describe how the compatibility required to exclude restricted-support mimicry changes with breadth and admissible genomic support. Because the shape of this surface also varies smoothly and reproducibly with *G* and *r*, we model the logistic parameters as functions *b*_0_(*G*, *r*, *C*_f_) and *k*(*G*, *r*, *C*_f_).

**Figure 5.**
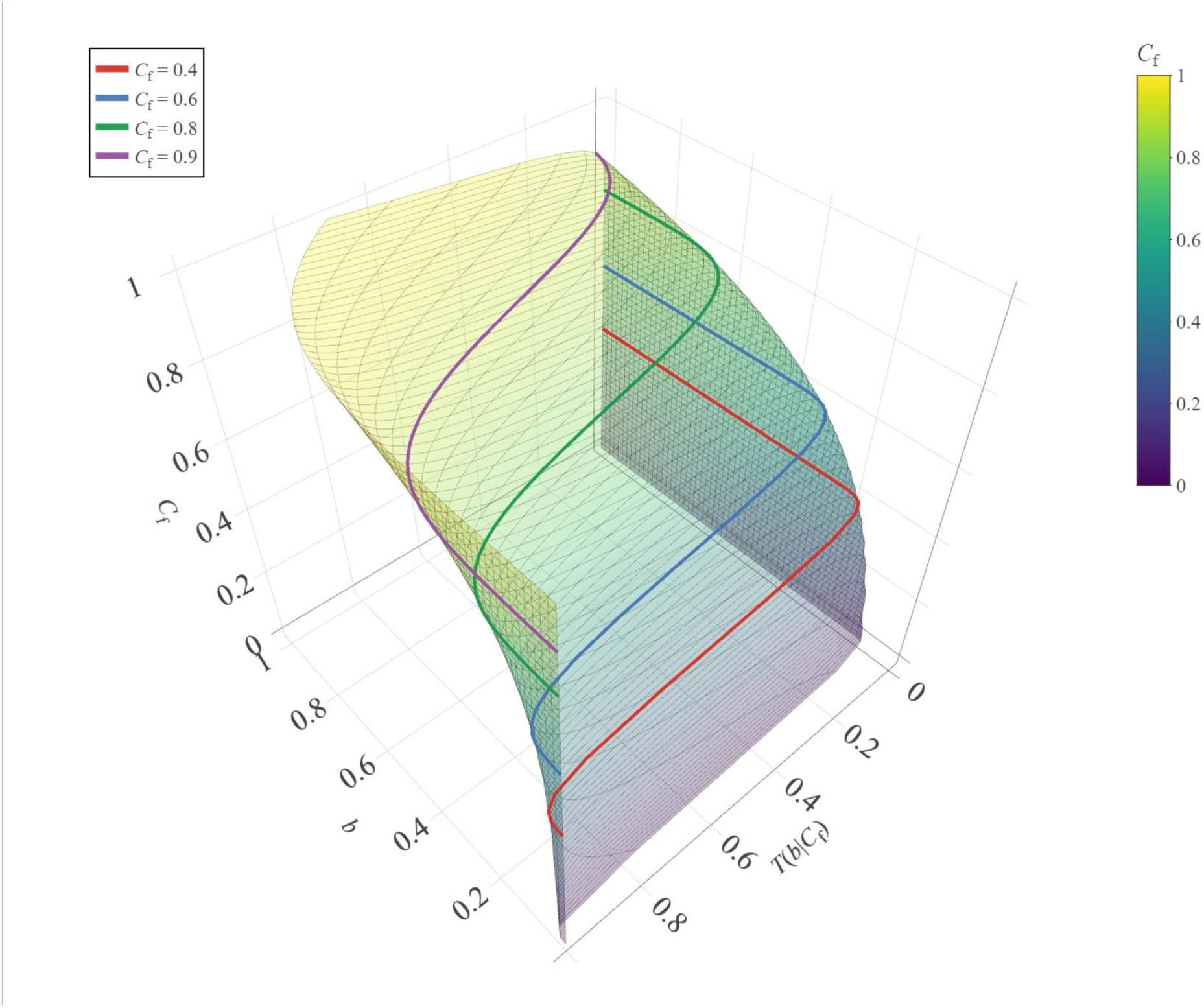
| Minimax compatibility surface for restricted-support alternatives. Semi-transparent surface showing the minimax compatibility boundary at *β*_0_ = 0.001 in restricted-support-fraction–breadth–dispersion-compatibility space. For fixed genome size *G* and read-length distribution *r*, the surface assigns to each combination of realised breadth *b* and restricted-support fraction *C*_f_ the threshold *T*(*b* | *C*_f_) above which mimicry by a *C*_f_-restricted alternative is excluded at 99.9% confidence. Values below the surface remain reproducible by restricted-support alternatives at the declared tolerance. Coloured curves show representative fixed-*C*_f_ slices at *C*_f_ = 0.4, 0.6, 0.8 and 0.9. Each slice follows the logistic boundary defined in Eq. (2), and together the slices reveal the smooth expansion of the admissible mimicry region as restricted genomic support increases. The surface is estimated from restricted-support Monte Carlo simulations and provides the compatibility geometry that is subsequently inverted over *C*_f_ to obtain the Metagenomic Alignment Validation Index (MAVI).

These functions were fitted to simulation-derived compatibility boundaries at *β*_0_ = 0.001 using 1,134 parameter settings from two structured calibration grids spanning *G* = 1–100 Mb, *r* = 1–6 kb and *C*_f_ = 0.3–0.9. Each setting was evaluated at 100 breadth points, with every boundary point estimated from 100,000 Monte Carlo realisations, yielding a high-precision representation of the empirical surface. The fitted parameterisation was validated against an additional 4,260 independently simulated settings covering *G* = 0.4–100 Mb, *r* = 0.6–15 kb and *C*_f_ = 0.3–0.9. Across validation runs, the analytically reconstructed surfaces reproduced the simulated boundaries with a mean pseudo-R^2^ of 0.97 ± 0.02 (Supplementary Note 5).

In practice, *G*, *r*, *b*_obs_ and Φ(*Z*_obs_) are observed for each candidate, leaving *C*_f_ as the only latent coordinate to be resolved on the minimax compatibility surface. For fixed *G* and *r*, each value of *C*_f_ defines a corresponding logistic boundary *T* (*b*∣ *G*, *r*,*C*_f_). We therefore evaluate this family of boundaries across *C*_f_ and identify the smallest restricted-support fraction whose boundary intersects the observed compatibility level at (*b*_obs_, Φ(*Z*_obs_)) by solving

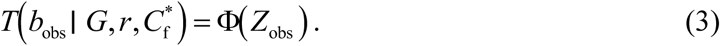

The value *C_t_*\* defines the Metagenomic Alignment Validation Index (MAVI), the one-dimensional genomic specialisation of the Generator Identifiability Envelope^18^ for metagenomic candidate validation (Supplementary Note 6). Specifically, *C_t_*\* is the smallest restricted-support fraction for which the observed breadth–compatibility coordinate remains statistically admissible at tolerance *β*_0_. Equivalently, MAVI is the minimum fraction of the candidate genome that a restricted-support explanation must access to mimic the observed compatibility with genome-wide random fragmentation.

Biologically, MAVI translates the observed dispersion under the stochastic fragmentation-and-survival process into a genomic support requirement. A high MAVI means that any restricted-support generator capable of mimicking the observed compatibility must access an unusually large fraction of the candidate genome, consistent with species-level genomic representation. Conversely, a low MAVI means that the same evidence remains reproducible from much more limited genomic support, leaving the species-level assignment unresolved. MAVI therefore provides a continuous, mechanistically interpretable bound on the minimum genomic support required by competing explanations.

#### Box 2

**– What MAVI tests in practical terms**

1. Take the mapped-read positions and lengths for each candidate species from the alignment output.
2. Compute the observed dispersion (*D*_obs_), calibrate it against the candidate-specific full-support random-fragmentation distribution of *D_s_*, and convert the result to a compatibility coordinate (*Z*, then Φ(*Z*)).
3. Reject candidates that are incompatible with random genome-wide fragmentation (Φ(*Z*) < *α*).
4. For the remaining candidates, invert the family of minimax compatibility boundaries over *C*_f_ to determine the smallest restricted genomic support that remains admissible.
5. Report the resulting MAVI for each candidate species.

<u>Verbatim example for MAVI = 0.8</u>

The observed alignment pattern has a MAVI of 0.80 at *β*_0_ = 0.001: **only explanations involving at least 80% shared genomic content of the candidate remain statistically admissible, with alternatives below this threshold excluded at ≥99.9% confidence**.

*This provides a direct statistical statement about the genomic extent required to explain the candidate alignment, rather than an inference reliant on heuristics or untestable assumptions*.

Although MAVI is continuous and does not rely on hard thresholds, its numerical scale has a direct biological interpretation. Comparative genomic studies show that shared gene or protein content is often on the order of 20–50% between genera within the same family, approximately 40–70% between species within the same genus, and more than 70–80% within the same species. These are not sharp taxonomic cut-offs, but broad empirical ranges that provide a simple interpretative guide, reflecting the general decrease in shared genomic content with increasing taxonomic distance^25,26^.

For example, a MAVI ≈ 0.20 for *Escherichia coli* indicates that the observed alignments remain compatible with a broad range of *Enterobacterales*, such as *Salmonella* or *Yersinia*. A MAVI around 0.50 restricts plausible alternatives to very closely related *Escherichia* species, such as *E. fergusonii* or *E. albertii*. Values above 0.8 indicate species-level confidence, where plausible alternatives are predominantly restricted to strain-level or near-clonal variants.

For practical interpretation and graphical representation, we therefore define conservative, lineage-agnostic ranges as a visual guide:

- **MAVI ≤ 0.30**: broadly compatible with family-level or broader taxonomic representation, including order, class or phylum.
- **0.30 < MAVI ≤ 0.50**: consistent with genus-level representation.
- **0.50 < MAVI < 0.75**: an intermediate region that naturally accommodates closely related species, species complexes and lineages with extensive shared genomic content, for which taxonomic boundaries are intrinsically blurred.
- **MAVI ≥ 0.75**: strong support for species-level presence.

These ranges are illustrative rather than prescriptive and can be adjusted for lineages with unusually high shared genomic content. In this way, plotting MAVI against breadth provides an immediate visual summary of taxonomic confidence within and across samples (Figure 6).

**Figure 6.**
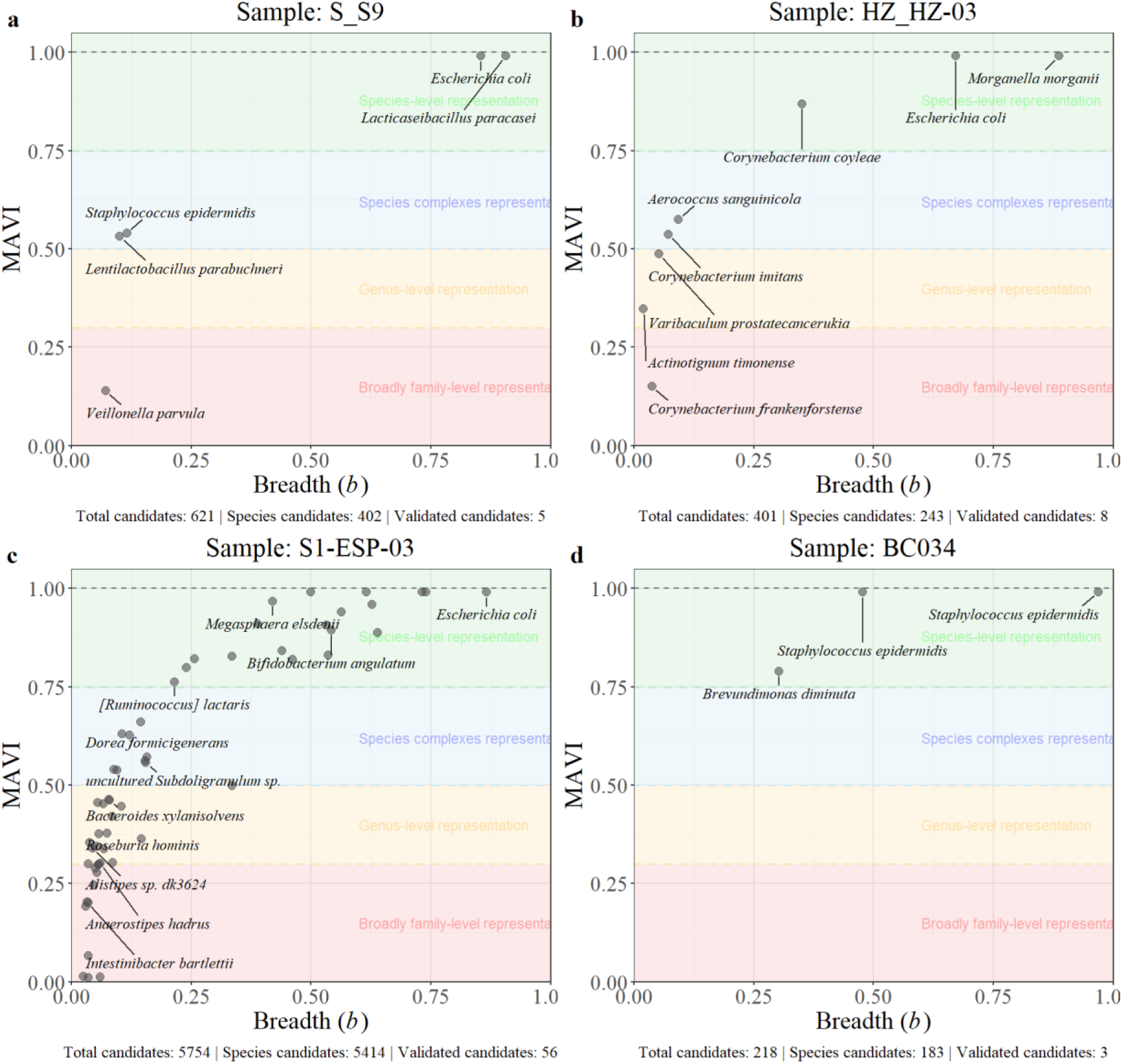
| MAVI validation landscapes across representative clinical metagenomic samples. Example outputs from four metagenomic samples: respiratory, S_S9; urine, HZ_HZ-03; stool, S1-ESP-03; and eye swab, BC034. Candidate reference genomes were taxonomically assigned using Kraken2, aligned with minimap2 and evaluated using MAVI. Each point represents a species-level candidate retained after the full-support compatibility gate, positioned according to its observed breadth *b* and corresponding MAVI. Coloured background bands are provided solely as visual guides to facilitate reading of the plot; they do not represent hard taxonomic boundaries and are not used in the MAVI calculation or inference. Counts below each panel report the total taxonomic candidates from Kraken2, species-level candidates and MAVI-validated candidates.

The analyses presented here use species-level reference genomes, reflecting the fact that species-level collections currently provide the most stable basis for reference-based metagenomic inference in the presence of strain diversity^27^. Within this framework, strain-specific variation is absorbed into the species-level interpretation. The same statistical principles can be applied at finer taxonomic resolution provided that high-quality strain-level reference genomes are available.

### 3.5 Minimum validated community explanation

MAVI evaluates candidate species individually, but validated candidates need not be supported by independent sequencing reads. The same recovered read may contribute evidence to several related species because of shared genomic sequence. Once candidate validation is complete, the read sets associated with the validated taxa induce a formal nested set structure under inclusion: identical read sets form equivalence classes, whereas strict subset relations define nested candidate explanations.

For each validated species, we tracked the reads contributing to its mapped evidence. This reveals a natural nesting among candidates. Some species are supported entirely by reads that are also represented by another validated species, whereas others contain private reads that cannot be accounted for by any other validated candidate. The latter are necessary components of any explanation that retains all validated sequencing evidence: removing one of these species leaves at least one recovered read unexplained. Candidates supported by identical read sets occupy the same primary read-support class, although their MAVI values may differ because the same reads can exhibit different genomic dispersion relative to different candidate genomes. MAVI therefore provides a secondary ordering among otherwise overlapping read-supported explanations.

We use this structure to identify the smallest number of species that collectively accounts for the reads represented by the MAVI-validated candidate set, which we term the minimum validated community explanation. Where several combinations explain the same read set equally well, the alternatives are retained rather than resolved arbitrarily. Species present in all minimum explanations are necessary components of the recovered community under the declared classification, alignment and reference regime.

This analysis in S10 (Figure 2b and Table 1) illustrates the full scope of MAVI, reducing 314 candidate calls comprising 254 species to a compact community-level explanation of the recovered sequencing evidence. Of all reads aligned to at least one candidate species, 97.63% were accounted for by candidates that passed the *α*-gate. Strikingly, 95.41% of the aligned read evidence was accounted for by only eleven necessary taxa with MAVI ≥ 0.01, with a further 0.34% contributed by competing taxas. An additional 1.88% of aligned reads could be explained only within the lower-MAVI (<0.01) residual layer. Of the total aligned read set, 1.13% comprised evidence private to individual residual necessary taxa, whereas the remaining 0.75% was shared among multiple admissible residual candidates. This uniquely required residual evidence, representing 1.13% of the aligned read set, was taxonomically concentrated: 29.3% of these reads were assigned to the genus *Avibacterium*, represented by a single taxon, while 61.0% belonged to genera already represented among the MAVI ≥ 0.01 taxa. The remaining 9.7% was distributed across taxa contributing only one or a few private reads. Thus, MAVI converts hundreds of candidate calls into a compact, explicitly structured explanation of the recovered community while preserving taxonomic ambiguity where the data cannot uniquely resolve it.

**Table 1.** | Read-support structure and minimum community explanation for S10. Taxa are classified by their role in explaining the read set after all-against-all candidate alignment and MAVI evaluation. Necessary taxa with MAVI ≥ 0.01 remain required after all alternative admissible explanations are considered. Competing taxa with MAVI ≥ 0.01 contribute read evidence that is required when considering taxa with MAVI ≥ 0.01, but their taxonomic identity is not uniquely required once alternative admissible explanations are allowed; in particular, a taxon may become unnecessary when taxa with MAVI < 0.01 are included in the minimum community explanation. Residual necessary taxa have MAVI < 0.01 but occur in every minimum validated community explanation, indicating that their read contribution is required for complete explanation despite insufficient support for stronger species-level validation. Redundant taxa with MAVI ≥ 0.01 are unnecessary even when considering only taxa with MAVI ≥ 0.01, because their read support is already explained by other taxa in that MAVI range. → denotes displacement by an alternative residual-community explanation. N is the number of species in each category. MAVI values are shown in parentheses and NCBI Taxonomy IDs in square brackets.

| Category | N | Taxa (MAVI) [Taxon ID] |
| --- | --- | --- |
| Necessary | 11 | *Haemophilus influenzae* ($>0.99$) [727]<br>*Streptococcus mitis* ($>0.99$) [28037]<br>*Streptococcus pseudopneumoniae* ($>0.99$) [257758]<br>*Haemophilus parainfluenzae* (0.90) [729]<br>*Streptococcus pneumoniae* (0.81) [1313]<br>*Streptococcus sanguinis* (0.73) [1305]<br>*Rothia dentocariosa* (0.69) [2047]<br>*Streptococcus salivarius* (0.67) [1304]<br>*Streptococcus oralis* (0.53) [1303]<br>*Granulicatella adiacens* (0.32) [46124]<br>*Veillonella parvula* (0.15) [29466] |
| Competing | 1 | *Neisseria sicca* (0.27) [490] → residual community |
| Residual necessary | 54 | *Avibacterium paragallinarum* ($<0.01$) [728] — 161 private reads<br>*Streptococcus koreensis* ($<0.01$) [2382163] — 158 private reads |
|  |  | *Streptococcus* sp. 11-4097 (<0.01) [2828286] — 34 private reads<br>*Streptococcus* sp. NPS 308 (<0.01) [1902136] — 31 private reads<br>*Streptococcus* sp. SN-1 (<0.01) [3074854] — 26 private reads<br>*Streptococcus gwangjuensis* (<0.01) [1433513] — 17 private reads<br>*Aggregatibacter aphrophilus* (<0.01) [732] — 13 private reads<br>*Streptococcus* sp. D7B5 (<0.01) [3038077] — 11 private reads<br>*Streptococcus* sp. HSISS3 (<0.01) [1316412] — 11 private reads<br>*Haemophilus parahaemolyticus* (<0.01) [735] — 5 private reads<br>*Streptococcus* sp. FSL W8-0197 (<0.01) [2975349] — 5 private reads<br>... |
| **Redundant** | 0 | None |

This analysis extends candidate-level validation into a community-level statement without requiring reconstruction of a unique latent community. It identifies which taxa are necessary to explain the recovered evidence, which are nested within other explanations, and where taxonomic ambiguity remains after validation.

## 4 Corroboration with independent clinical interpretation

For evaluation, we examined eight previously published long-read metagenomic datasets. Six comprised 280 clinical samples in total: Charalampous, et al. ^23^ (83 samples), Bellankimath, et al. ^28^ (59 samples), Alcolea-Medina, et al. ^29^ (66 samples), Campos-Madueno, et al. ^30^ (25 samples), Jing, et al. ^31^ (25 samples), and Nielsen, et al. ^32^ (22 samples). Experimental validation datasets included in these studies—such as mock communities, organism-spiked matrices, serial-dilution series and limit-of-detection experiments—were also analysed separately from the clinical cohorts. We additionally included a wastewater study by Che, et al. ^33^ (11 samples) and a single-organism sample from Rautiainen and Marschall ^34^, giving a total of 643 samples.

The published interpretations were treated as independent reference points because they were informed by combinations of culture, expert assessment and, in complex cases, targeted molecular assays. We used them to assess whether MAVI recovers, from metagenomic data alone, the same candidate-level biological interpretations reached through substantially more involved validation workflows, and whether it provides additional structure where the original evidence remained heuristic or equivocal.

Across the clinical respiratory samples analysed by Charalampous, et al. ^23^, MAVI consistently validated the organisms ultimately supported by the original study in every case (Supplementary Data 1), including those corroborated by culture or targeted molecular assays. Importantly, where the original analysis required expert judgement or follow-up molecular testing to resolve sparse and uneven metagenomic evidence, MAVI reached the same conclusions using dispersion-based evidence alone. For example, in sample S9 (Figure 6a), culture reported a mixed infection with *Pseudomonas aeruginosa* and *Escherichia coli*. Metagenomic sequencing did not support *P. aeruginosa*, but returned a broader set of possible candidates, with *E. coli* as the leading candidate, leaving the species-level interpretation unresolved from sequencing alone. Follow-up qPCR excluded *P. aeruginosa* and corroborated *E. coli*. MAVI reached the same conclusion directly from the metagenomic alignments, identifying *E. coli* as the dominant genome-wide signal and separating it from lower-confidence background candidates. Across these cases, MAVI was concordant with the organism-level conclusions supported by independent clinical or molecular evidence.

Across the urinary tract infection samples analysed by Bellankimath, et al. ^28^, MAVI was fully concordant with the organisms reported in the original study (Supplementary Data 1). In the original workflow, metagenomic detections were interpreted primarily as binary pathogen calls against routine culture, with additional detections subsequently assessed using Vivalytic or pathogen-specific PCR. The primary distinction therefore lies in interpretation rather than detection: MAVI quantifies the extent of genome-wide representation underlying each call. In polymicrobial samples, MAVI further resolves relative genomic representation by distinguishing dominant genome-wide signals from marginal candidates. MAVI also provides information on taxa beyond those reported in the original analysis, enriching sample-level interpretation. For example, in sample HZ-03, MAVI identified *Varibaculum spp*. with genus-level genomic confidence (Figure 6b), revealing a clinically relevant taxon that was present in the metagenomic output but not carried forward in the original corroboration workflow because it fell outside the predefined validation set.

Across the stool metagenomic datasets analysed by Campos-Madueno, et al. ^30^, MAVI validated the bacterial species reported as present by the original study across the cohort (Supplementary Data 1). The original study pursued a different analytical objective, focusing on selected bacteria and their plasmid or chromosomal genetic environments; concordance with its reported species therefore provides corroboration rather than a direct validation of MAVI across the full metagenome. These datasets are nevertheless particularly informative because each sample contains thousands of candidate taxa, for which exhaustive candidate-by-candidate corroboration would require an impractical number of orthogonal analyses. MAVI instead evaluated the complete candidate set directly from the alignment output, recovering the reported species while assigning an explicit identifiability bound to every candidate despite extensive plasmid- and chromosome-mediated genomic sharing (Figure 6c). A representative example is stool sample S1-ESP-03, which contained more than 5,000 candidate taxa, with similar magnitudes observed across the other samples. MAVI distinguished the reported species from thousands of alternative taxa, including many with substantial breadth or read counts that conventional thresholds could have promoted as valid detections or left inconclusive. Overall, MAVI reduced thousands of otherwise unresolved candidates to a few dozen candidate species with explicit genomic-support bounds.

Across the ocular surface cohort analysed by Jing, et al. ^31^, MAVI corroborated the dominant species reported in the original study and additionally resolved low-abundance candidates that were not individually interpreted in its read-count-based reporting framework (Supplementary Data 1). This cohort provides a particularly stringent test because the ocular surface is a naturally paucibacterial environment, where sparse and low-abundance microbial evidence is expected rather than exceptional. The original analysis classified bacterial detections using a threshold of more than three unique reads and evaluated agreement primarily through the dominant flora, leaving the genomic extent underlying lower-abundance detections unresolved. MAVI detected *Citrobacter koseri* in four independent samples (Supplementary Data 1). In addition to the occurrence reported in the original study, two low-breadth alignments occupied the species-complex interpretative range, with MAVI values of 0.61 and 0.66 (Supplementary Data 1), showing that competing explanations would require access to at least 61% and 66% of the candidate genome, respectively. Similarly, *Brevundimonas* was present in the genus-level composition reported by the original study, but its species-level interpretation remained unresolved. MAVI identified *Brevundimonas diminuta* in a single low-breadth sample and placed its genomic support within the species-complex confidence range (Figure 6d). These results show that MAVI can recover interpretable genomic-support bounds even in a naturally low-biomass setting, where fixed read-count thresholds are particularly prone to discarding informative low-abundance signals or leaving them unresolved.

### Box 3

**– Clinical Vignette: resolving ambiguity in sparse metagenomic data**

A clear example is sample S10 from Charalampous, et al. ^23^. In this case, routine culture did not yield a definitive pathogen consistent with the final clinical interpretation due to heavy mixed growth. Metagenomic sequencing suggested the high prevalence of *Streptococcus pneumoniae* and *Haemophilus influenzae*, but these signals were embedded within a large set of more than 50 candidate taxa, occupying an ambiguous breadth–read count regime. This ambiguity precluded confident species-level assignment based on sequencing alone and led the sample to be classified as inconclusive.

Targeted molecular assays were subsequently performed: *ply* gene qPCR for *Streptococcus pneumoniae* and *hpd*/*ompP6* gene qPCR for *Haemophilus influenzae*, both of which returned positive results, confirming S10 as a true mixed infection. Notably, MAVI reached the same validation directly from metagenomic data alone: Figure 2b shows the compression of the complex candidate space to strong species-level genomic support, while the minimum validated community explanation in Table 1 identifies both organisms as necessary components of the recovered sequencing evidence. Together, these analyses provide strong species-level genomic confidence for both organisms without reliance on culture or follow-up molecular validation. MAVI provides a principled, statistically explicit and physically grounded characterisation of taxon presence, yielding traceable, auditable and fully reproducible evidence that can support decision-making in complex or contested clinical contexts.

Across the plasma cell-free DNA metagenomic datasets analysed by Nielsen, et al. ^32^, MAVI identified the bacterial species reported as present by the original study (Supplementary Data 1). As expected for bloodstream cfDNA sequencing, genomic support remained extremely sparse and identifiability bounds were uniformly low; nevertheless, the dispersion structure of these trace-level signals was concordant with the organisms reported in the original analysis. This illustrates that MAVI remains consistent with the published interpretation even in the extreme low-coverage regime characteristic of plasma cfDNA metagenomics.

An instructive proof of concept is provided by the single-organism dataset used by Rautiainen and Marschall ^34^, consisting of *Escherichia coli* K-12 PacBio HiFi reads that we processed through the same long-read metagenomic pipeline. Despite originating from a monoculture, the pipeline produced about a dozen of taxonomic candidates, including *Escherichia albertii* and *Shigella spp*. Using dispersion-based validation alone, MAVI rejected all alternative candidates and uniquely validated *E. coli* as the only taxon exhibiting genome-wide representation (Extended Data Fig. 1). Although strain-level inference is not a primary aim of this study, MAVI further narrowed support to three closely related *E. coli* strains, including K-12, consistent with the known ground truth (Supplementary Data 1). This example highlights how MAVI can resolve taxonomic ambiguity arising purely from shared genomic regions, even in the absence of biological complexity, and provides a cross-platform proof of concept beyond Nanopore sequencing.

A complementary proof of concept is provided by the datasets analysed by Che, et al. ^33^, encompassing influent, activated sludge and effluent samples from three wastewater treatment plants. Across all eleven datasets, MAVI validated the species reported as present by the original study (Supplementary Data 1). Notably, MAVI reduced more than ten thousand initial taxonomic candidates per sample to approximately one hundred supported candidates, with explicit species- or genus-level genomic-support bounds. These samples represent an extreme regime characterised by high microbial complexity, extensive horizontal gene transfer and pervasive mobile genetic elements. Despite this, MAVI produced clean and unambiguous validation outcomes, rejecting alternative taxa whose apparent breadth and read counts could otherwise render them inconclusive or falsely acceptable under ad hoc threshold-based criteria. This demonstrates that the framework collapses taxonomic uncertainty by multiple orders of magnitude even under highly heterogeneous and recombinogenic conditions.

Across the respiratory samples analysed by Alcolea-Medina, et al. ^29^, MAVI was concordant with the taxa reported in the original study (Supplementary Data 1). However, bacterial recovery was sparse throughout the cohort. Outside the dominant taxa, most candidates therefore received low genomic-support bounds. More importantly, unlike the other respiratory cohorts examined here, the MAVI response repeatedly collapsed into a small number of discrete, near-binary states. Such a pattern may occur in individual samples through biological variation. However, its systematic recurrence across the cohort is unusual and suggests that stochastic dispersion may have been truncated during wet-lab processing. This does not call into question the taxa validated from the surviving material, but it may limit how fully the wider microbial community can be interpreted.

Within the MAVI framework, preservation or loss of this stochasticity leaves an observable signature. MAVI is derived from the physical dispersion law induced by stochastic genome fragmentation and fragment survival. Each candidate is therefore placed on a coordinate reflecting how much of the expected stochastic structure remains in the observed alignments. The fragmentation mechanism becomes empirically interrogable rather than merely assumed. At cohort level, the normalised generator-level dispersion entropy introduced by Márquez and Silva-Toro ^19^ makes this structure explicit by summarising the diversity of candidate compatibility states. We formalise this diagnostic perspective for metagenomic workflows below.

### 4.1 Cohort-level stochastic structure and entropy-based assessment

MAVI is built on a single central premise formalised by Lander and Waterman ^20^: genomic fragments are sampled randomly across the genome. In the absence of selective enrichment or depletion, the resulting sequencing reads therefore represent the stochastic survival of genomic fragments through sample preparation and sequencing. Under this regime, genuine genome-wide presence is not expected to collapse systematically into a small number of discrete alignment states. Instead, it generates a continuous dispersion structure shaped by random fragmentation, conserved regions and shared homology. MAVI makes this stochastic geometry explicit by converting each candidate’s observed dispersion into the candidate-specific compatibility coordinate Φ(*Z*). The cohort-level distribution of these coordinates thereby renders the retained stochastic structure directly observable rather than implicit.

The stochastic structure of the workflow extends beyond any individual taxon or sample and is therefore assessed at the cohort level. We quantify it using the normalised generator-level dispersion entropy introduced by Márquez and Silva-Toro ^19^ for Boolean coverage processes (Supplementary Note 7). The entropy is calculated from the pooled distribution of candidate-specific compatibility coordinates, Φ(*Z*), across samples within a cohort. A distribution occupying many compatibility states produces high entropy, whereas concentration into a small number of states produces low entropy. The resulting value therefore describes how much stochastic variability is preserved by the workflow across the cohort.

This entropy is not an absolute measure of biological diversity. It characterises how metagenomic evidence is distributed under a particular combination of biological context and analytical workflow. Different sample types may therefore occupy different characteristic entropy regimes because they correspond to different underlying generators and recovery structures. Entropy values should consequently be interpreted within matched biological contexts rather than compared indiscriminately across sample types. Differences between cohorts of the same sample type can then reveal truncation, selective loss or other workflow-dependent distortions of the expected stochastic structure.

Across the datasets analysed here, cohort-level entropy followed a coherent sample-type ordering (Extended Data Fig. 2), with stool^30^ and respiratory^23^ samples exhibiting the highest values (0.675 and 0.699), followed by urine^28^ (0.608) and ocular-surface^31^ samples (0.497). This ordering was consistent with the expected generator structure of each sample type, including its biological origin, biomass-recovery profile and the complexity of the recoverable microbial community. Each cohort retained a broad distribution of compatible Φ(*Z*) states shaped by that underlying structure. These results support the theoretical expectation developed above: in the absence of selective enrichment or depletion, entropy should occupy a characteristic and partially reproducible regime for a given sample type, despite variation in the particular samples represented within each cohort.

The dataset generated using the workflow of Alcolea-Medina, et al. ^29^ provides a clear and informative contrast. Based on the sample-type ordering observed across the study, these respiratory samples would be expected to group with the other respiratory cohorts and retain their position relative to stool, urine and ocular-surface samples. Instead, candidate taxa repeatedly collapsed into a small number of discrete compatibility states across the cohort. This produced substantially reduced normalised generator-level dispersion entropy (0.5) and a pronounced shift towards near-binary structure (compare Extended Data Fig. 2 and Extended Data Fig. 3). The resulting entropy regime was closer to the ocular-surface cohort than to the broader respiratory pattern.

This departure from the expected regime is inconsistent with unbiased stochastic fragmentation and points to substantial upstream truncation of fragment survival. The analysis cannot identify the responsible preparation step. The effect could arise during host depletion, nuclease treatment, nucleic-acid conversion, barcoding, or through interactions among these stages. Under such conditions, MAVI can still validate taxa represented in the surviving genomic material. Inference about the full underlying microbial community nevertheless becomes ill-posed because the preparation process dominates which fragments remain observable. Consequently, non-recovery in this cohort should be interpreted cautiously, as the reduced stochastic dispersion is consistent with greater workflow-dependent loss of recoverable genomic material relative to the other datasets examined.

Crucially, this failure mode is not silent within the MAVI framework: loss of the expected stochastic dispersion is directly observable through cohort-level entropy collapse. The physical sampling premise underlying MAVI is therefore empirically testable and falsifiable rather than merely assumed.

## 5 Conclusion

This study establishes metagenomic validation as an inference problem governed by the physical processes that generate sequencing data. Random genome fragmentation and fragment survival produce an observable dispersion law, while fragment loss truncates the genomic support available for inference. We formally show that, because the geometry of this truncation is unobserved, the same mapped reads may be compatible with either genome-wide presence or restricted genomic support. Species presence is therefore generally non-identifiable from alignment evidence alone. To resolve what remains identifiable, we invert the physical dispersion law over the admissible class of restricted-support alternatives under a minimax criterion. This inversion defines the Metagenomic Alignment Validation Index (MAVI). MAVI returns a mechanistically interpretable lower bound: the minimum fraction of the candidate genome that any competing explanation must access to reproduce the observed evidence.

Across clinical, environmental and artificial-control samples, MAVI recovered independently supported taxa while assigning an explicit genomic-support bound to every candidate. It distinguished candidates requiring broad genomic support from those reproducible under restricted support across sparse, polymicrobial and highly complex metagenomic regimes. These results demonstrate that low or uneven coverage does not automatically invalidate metagenomic evidence. Instead, it determines how strongly restricted-support explanations can be excluded. When the available evidence is insufficient for certification, MAVI retains that ambiguity explicitly rather than replacing it with a threshold-based decision.

The same mechanistic formulation makes the underlying physical premise empirically testable through cohort-level entropy. It also provides a computationally lightweight validation layer that can be applied across thousands of candidates without requiring candidate-specific marker selection or exhaustive orthogonal confirmation. In the datasets examined here, MAVI reduced taxonomic ambiguity by multiple orders of magnitude while preserving continuous, biologically interpretable support statements. Metagenomic detection and validation must therefore be distinguished: detection establishes sequence compatibility, whereas validation asks whether restricted-support explanations can be excluded at a declared level of confidence.

## 6 Materials and Methods

### 6.1 Taxonomic profiling

Raw sequencing reads were first subjected to host-read removal using Hostile (v2.0.0), with alignment to the human reference genome GRCh38 performed using minimap2. The resulting host-depleted reads were taxonomically classified using Kraken2 (v2.1.3) with the standard database downloaded in December 2025, without imposing a confidence threshold. Per-read classification files and summary reports were generated, and reads assigned to candidate taxa were extracted using KrakenTools (extract_kraken_reads.py). Candidate taxa were subsequently re-examined by mapping the extracted reads to their corresponding species-level RefSeq genomes using minimap2 (v2.28) with the map-ont preset. Alignment files were processed using SAMtools (v1.21). These alignments formed the basis for all dispersion, coverage-breadth and Φ(Z) calculations described in the main text.

### 6.2 Numerical procedures and implementation details

The numerical implementation followed the MAVI procedure defined in the main text and detailed in the Supplementary Information. Candidate-specific inputs were obtained from the minimap2 alignments and included genome size, mapped-read count, observed breadth and dispersion, and the empirical read-length distribution. Dispersion was calculated using a fixed 1,000-bp gap threshold, with identical merging rules applied to observed and simulated alignments. Candidate-specific null distributions were estimated using 1,000 Monte Carlo replicates, and MAVI values were obtained by numerical inversion of the pre-trained restricted-support boundaries. Monte Carlo null-variance collapse cases, in which the simulated null dispersion had effectively zero variance, were retained and passed directly to MAVI inversion, without assigning rejection or acceptance at the *α*-gate stage, as described in Supplementary Note 1.2.

MAVI was implemented in Python using NumPy, SciPy and pysam and applied as a single-node post-hoc validation layer to minimap2 alignments. The complete workflow, including SAMtools-based preprocessing, was executed using 4 CPU cores and 16 GB RAM per sample. The inferential core required 2 CPU cores and 8 GB RAM and completed in seconds to minutes per sample. Samples were processed concurrently under independent single-node Slurm allocations.

The computational logic, statistical construction, parameterisation and procedures required to reproduce MAVI are described in full in Supplementary Notes 1-7. Source code and analysis scripts required to reproduce the reported analyses will be deposited in a public repository upon publication.

All model training and validation, read mapping, and dispersion calculations were performed on the Birmingham Environment for Academic Research (BEAR) high-performance computing service at the University of Birmingham.

### 6.3 Reference datasets

All sequencing datasets were obtained from publicly available studies (PRJEB30781 [respiratory], PRJEB83412 [urine], PRJNA890545 [ocular surface], PRJNA931814 [stool], PRJNA1108520 [blood], PRJEB61294 [respiratory], PRJNA479723 [wastewater] and SRR10971019 [*E. coli*]). Reported taxonomic interpretations were taken from the authors’ original conclusions, which were based on their own validation criteria, including culture confirmation, assembly-based evidence, marker thresholds and other study-specific methods. These interpretations were used only as external references against which to compare the behaviour of our dispersion-based framework.

### 6.4 Minimum validated community explanation

Original read identifiers were retained through taxonomic classification, candidate-read extraction and reference alignment. For community-level analysis, reads were independently aligned against each candidate species reference using minimap2. All reported alignments were retained, without additional MAPQ, query-coverage or sequence-identity filtering, so that alternative candidate explanations remained represented. Duplicate or supplementary alignments from the same read to the same candidate were collapsed to a single read identifier.

For each sample, the unique read identifiers contributing to each candidate taxon were collected to define its read-support set. Candidates with a successful post-*α* MAVI outcome were retained as admissible explanations, whereas α-rejected candidates and candidates without a successful MAVI evaluation were excluded from the validated-community optimisation. Community structure was then resolved using exact minimum set-cover optimisation. First, the smallest set of taxa with MAVI ≥ 0.01 whose combined read-support sets accounted for the read evidence represented within that MAVI range was identified.

Candidate membership across equally minimal solutions was evaluated to distinguish taxa that were necessary, competing or redundant within this layer. The optimisation was then repeated after admitting taxa with MAVI < 0.01, over the complete read set represented by all *α*-admissible candidates. This second step determined whether taxa with MAVI ≥ 0.01 remained necessary after lower-MAVI alternative explanations were allowed and identified residual taxa required to complete the minimum validated community explanation. MAVI values were not used as weights in either optimisation.

For biological interpretation, we quantified the fraction of aligned read evidence represented by *α*-admissible candidates, by taxa with MAVI ≥ 0.01, and by necessary taxa within that range. Read evidence that could only be explained after admitting candidates with MAVI < 0.01 was retained as a residual layer. Taxa with MAVI < 0.01 that occurred in every minimum validated community explanation were classified as residual necessary taxa. Alternative taxonomic explanations were preserved rather than resolved by an arbitrary choice between equally admissible minimum solutions. Full mathematical definitions are provided in Supplementary Note 8.

### 6.5 Generator-level dispersion entropy

Generator-level dispersion entropy was calculated from the distribution of candidate-specific Φ(*Z*) values. Candidates were pooled across all samples within each experimental group, and only those passing the compatibility gate, Φ(*Z*) ≥ *α* = 0.001, were included. The interval [α,1] was partitioned into *B*=25 equal-width bins. For bin probabilities *p*_j_, estimated as the proportion of eligible candidates within each bin, entropy was calculated as

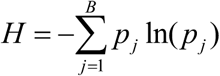

and normalised as

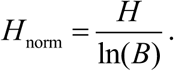

Groups containing fewer than 20 eligible candidates were designated insufficient for entropy estimation. Full mathematical definitions are provided in Supplementary Note 7.

## Data Availability

All data generated in the present study are contained in the manuscript and Supplementary Information. All source sequencing datasets are publicly available under the accession numbers listed in the manuscript.

## 7 Acknowledgments

The authors thank the University of Birmingham for providing access to the BlueBEAR high-performance computing service and its technical staff for supporting the computational work undertaken in this study.

## 8 Author contributions

MB identified the limitations in current validation metrics. MB and DAM conceived the study. DAM developed the minimax identifiability framework underlying MAVI and formalised stochastic fragment loss as a generator-level inferential signal for metagenomic validation. DAM and FS-T derived and validated the model. DAM and MB analysed the data and conducted the proof-of-concept analyses. MB led the microbiological interpretation. DAM implemented MAVI and drafted the manuscript. All authors contributed to data interpretation and revision of the manuscript.

## Extended data

**Extended Data Fig. 1.**
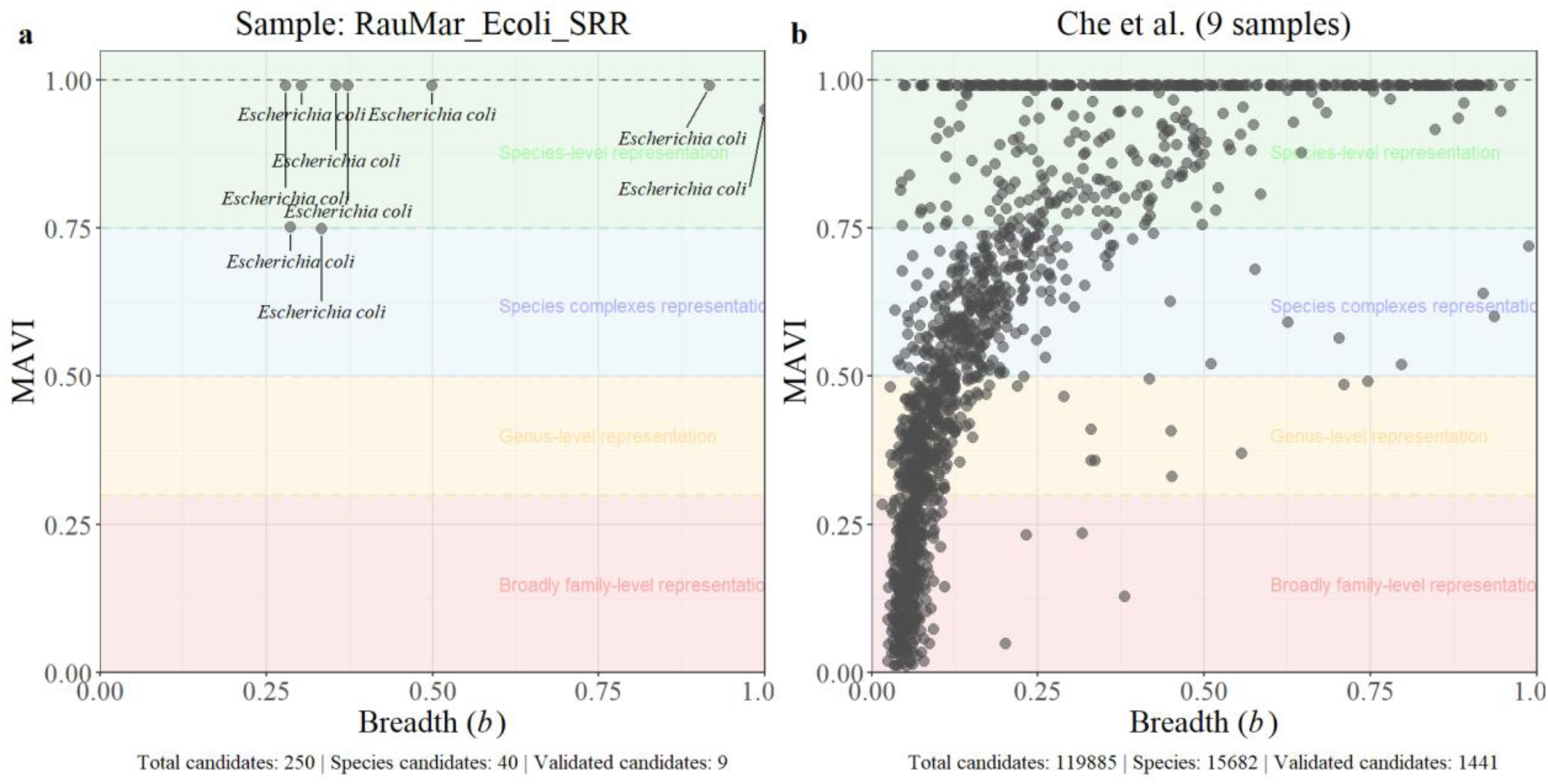
| MAVI validation landscapes under controlled and highly heterogeneous conditions. (a) Breadth–MAVI landscape for the single-organism *Escherichia coli* K-12 dataset used by Rautiainen and Marschall^34^. (b) Breadth–MAVI landscape across representative wastewater samples from Che et al.^33^. Each point represents a candidate reference genome evaluated independently by MAVI. Coloured background bands are included solely as interpretive guides; they do not define hard taxonomic boundaries and are not used to calculate MAVI.

**Extended Data Fig. 2.**
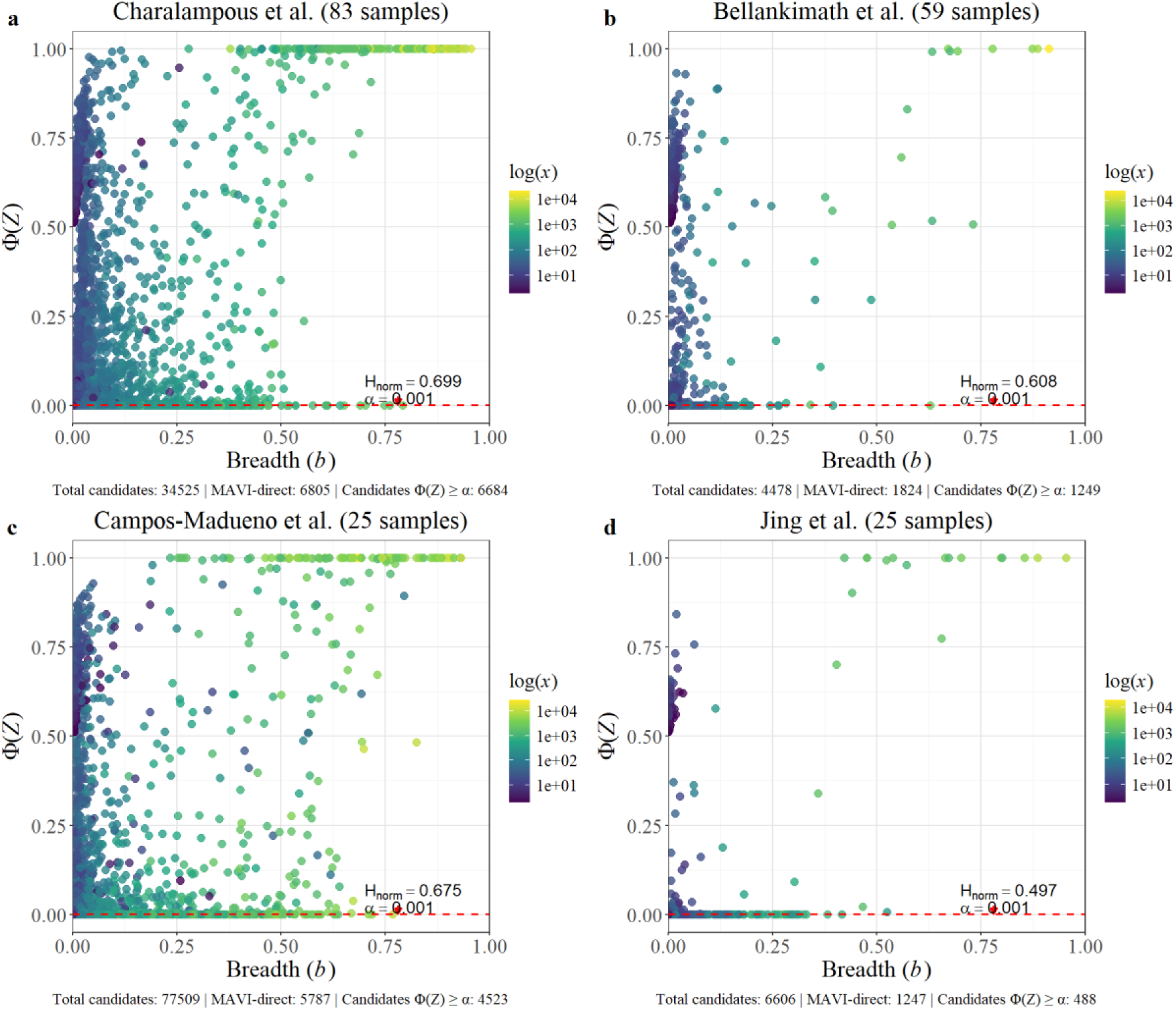
| Cohort-level stochastic dispersion structure across independent clinical metagenomic studies. Breadth– Φ(Z) distributions for all candidate reference genomes across the respiratory (Charalampous et al.^23^), urinary (Bellankimath et al.^28^), stool (Campos-Madueno et al.^30^) and ocular-surface (Jing et al.^31^) cohorts. Each point represents a candidate genome evaluated independently by MAVI and is coloured by the logarithm of the number of mapped reads (*x*). Red dashed lines indicate the dispersion-compatibility threshold (*α* = 0.001). The associated normalised generator-level dispersion entropy (*H*_norm_) is calculated from the pooled Φ(Z) distribution of candidates satisfying Φ(Z) ≥ *α* and summarises the retained stochastic dispersion structure within each cohort.

**Extended Data Fig. 3.**
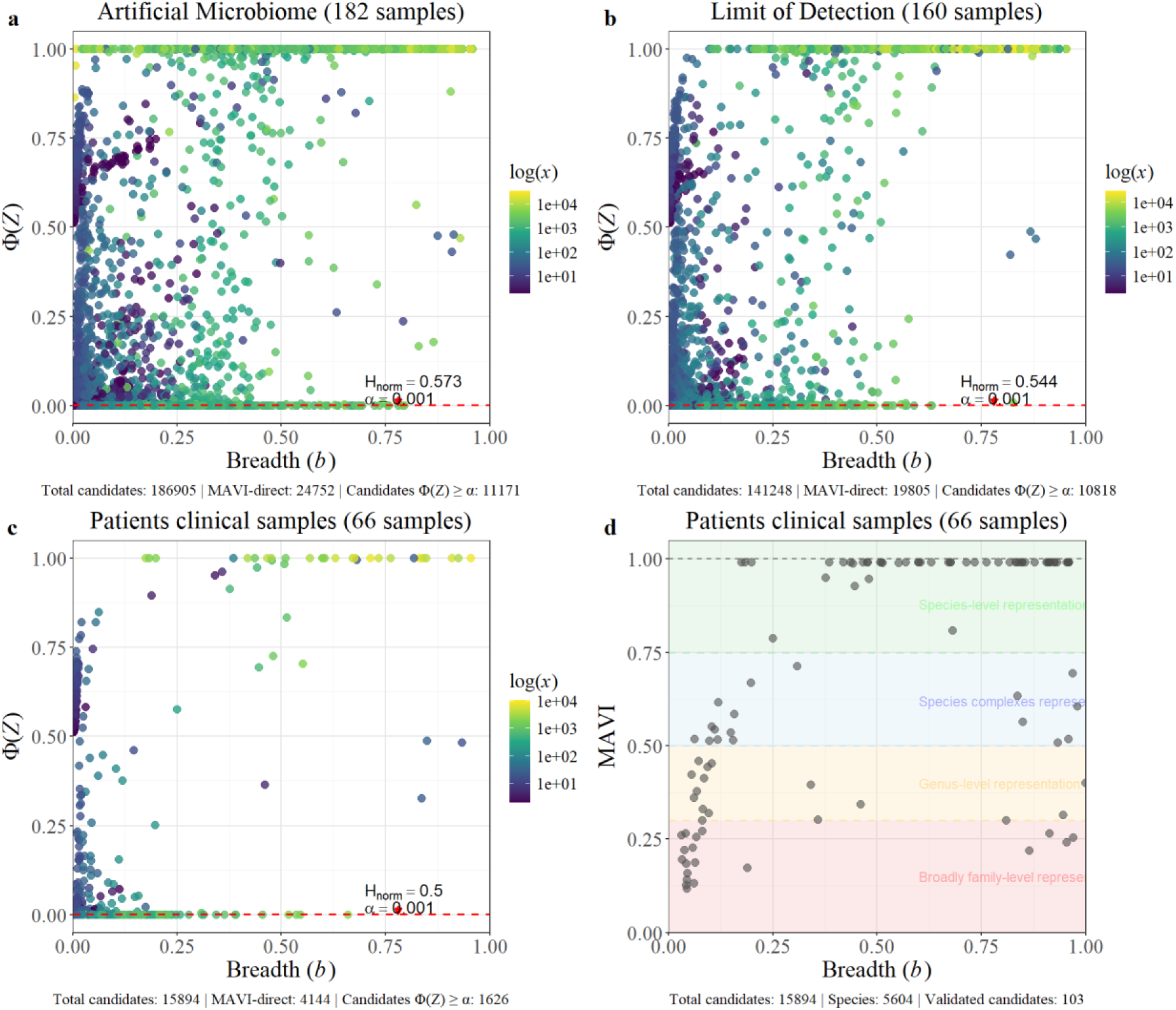
| Cohort-level stochastic dispersion structure across artificial, limit-of-detection and clinical sample sets. Breadth– Φ(Z) distributions for datasets reported in Alcolea-Medina et al.^29^. Each point represents a candidate genome evaluated independently by MAVI and is coloured by the logarithm of the number of mapped reads (*x*). Red dashed lines indicate the dispersion-compatibility threshold (*α* = 0.001). The associated normalised generator-level dispersion entropy (*H*_norm_) is calculated from the pooled Φ(Z) distribution of candidates satisfying Φ(Z) ≥ *α* and summarises the retained stochastic dispersion structure within each cohort. The lower right panel shows the distribution of patient-derived candidates annotated by MAVI.

## References

1 Diao, Z. et al. Assessing the Quality of Metagenomic Next-Generation Sequencing for Pathogen Detection in Lower Respiratory Infections. Clin. Chem. 69, 1038–1049 (2023). 10.1093/clinchem/hvad072

2 Liu, D. et al. Multicenter assessment of shotgun metagenomics for pathogen detection. eBioMedicine 74 (2021). 10.1016/j.ebiom.2021.103649

3 Chen, L. et al. Short- and long-read metagenomics expand individualized structural variations in gut microbiomes. Nature Communications 13, 3175 (2022). 10.1038/s41467-022-30857-9

4 Agustinho, D. P. et al. Unveiling microbial diversity: harnessing long-read sequencing technology. Nat. Methods 21, 954–966 (2024). 10.1038/s41592-024-02262-1

5 Kim, C., Pongpanich, M. & Porntaveetus, T. Unraveling metagenomics through long-read sequencing: a comprehensive review. Journal of Translational Medicine 22, 111 (2024). 10.1186/s12967-024-04917-1

6 Portik, D. M., Brown, C. T. & Pierce-Ward, N. T. Evaluation of taxonomic classification and profiling methods for long-read shotgun metagenomic sequencing datasets. BMC Bioinformatics 23, 541 (2022). 10.1186/s12859-022-05103-0

7 Wood, D. E., Lu, J. & Langmead, B. Improved metagenomic analysis with Kraken 2. Genome Biology 20, 257 (2019). 10.1186/s13059-019-1891-0

8 Breitwieser, F. P., Baker, D. N. & Salzberg, S. L. KrakenUniq: confident and fast metagenomics classification using unique k-mer counts. Genome Biology 19, 198 (2018). 10.1186/s13059-018-1568-0

9 Kim, D., Song, L., Breitwieser, F. P. & Salzberg, S. L. Centrifuge: rapid and sensitive classification of metagenomic sequences. Genome Res. 26, 1721–1729 (2016). 10.1101/gr.210641.116

10 Ounit, R., Wanamaker, S., Close, T. J. & Lonardi, S. CLARK: fast and accurate classification of metagenomic and genomic sequences using discriminative k-mers. BMC Genomics 16, 236 (2015). 10.1186/s12864-015-1419-2

11 Li, H. Minimap2: pairwise alignment for nucleotide sequences. Bioinformatics 34, 3094–3100 (2018). 10.1093/bioinformatics/bty191

12 Sun, Z. et al. Removal of false positives in metagenomics-based taxonomy profiling via targeting Type IIB restriction sites. Nature Communications 14, 5321 (2023). 10.1038/s41467-023-41099-8

13 Franzosa, E. A. et al. Identifying personal microbiomes using metagenomic codes. PNAS 112, E2930–E2938 (2015). doi:10.1073/pnas.1423854112

14 Bowers, R. M. et al. Minimum information about a single amplified genome (MISAG) and a metagenome-assembled genome (MIMAG) of bacteria and archaea. Nat. Biotechnol. 35, 725–731 (2017). 10.1038/nbt.3893

15 Koslicki, D., White, S., Ma, C. & Novikov, A. YACHT: an ANI-based statistical test to detect microbial presence/absence in a metagenomic sample. Bioinformatics 40 (2024). 10.1093/bioinformatics/btae047

16 Olm, M. R. et al. Robust variation in infant gut microbiome assembly across a spectrum of lifestyles. Science 376, 1220–1223 (2022). 10.1126/science.abj2972

17 Sanguineti, D., Zampieri, G., Treu, L. & Campanaro, S. Metapresence: a tool for accurate species detection in metagenomics based on the genome-wide distribution of mapping reads. mSystems 9, e00213–00224 (2024). doi:10.1128/msystems.00213-24

18 Márquez, D. A. & Silva-Toro, F. Generator Identifiability Envelope: Minimax support-fraction inference for Boolean random closed sets under hidden support restriction. (2026). 10.5281/zenodo.22884693

19 Márquez, D. A. & Silva-Toro, F. Generator-level dispersion laws for Boolean random closed sets under partial observation. (2026). 10.5281/zenodo.22884521

20 Lander, E. S. & Waterman, M. S. Genomic mapping by fingerprinting random clones: A mathematical analysis. Genomics 2, 231–239 (1988). 10.1016/0888-7543(88)90007-9

21 Hammersley, J. M. & Handscomb, D. C. Monte Carlo Methods. (Springer Dordrecht, 1964).

22 Manly, B. Randomization, Bootstrap and Monte Carlo Methods in Biology. (2007).

23 Charalampous, T. et al. Nanopore metagenomics enables rapid clinical diagnosis of bacterial lower respiratory infection. Nat. Biotechnol. 37, 783–792 (2019). 10.1038/s41587-019-0156-5

24 Karlin, S. & Brendel, V. Patchiness and correlations in DNA sequences. Science 259, 677–680 (1993). 10.1126/science.8430316

25 Tu, Q. & Lin, L. Gene content dissimilarity for subclassification of highly similar microbial strains. BMC Genomics 17, 647 (2016). 10.1186/s12864-016-2991-9

26 Qin, Q.-L. et al. A Proposed Genus Boundary for the Prokaryotes Based on Genomic Insights. J. Bacteriol. 196, 2210–2215 (2014). doi:10.1128/jb.01688-14

27 Konstantinidis, K. T., Ramette, A. & Tiedje, J. M. The bacterial species definition in the genomic era. Philos Trans R Soc Lond B Biol Sci 361, 1929–1940 (2006). 10.1098/rstb.2006.1920

28 Bellankimath, A. B. et al. Metagenomic sequencing enables accurate pathogen and antimicrobial susceptibility profiling in complicated UTIs in approximately four hours. Nature Communications (2025). 10.1038/s41467-025-66865-8

29 Alcolea-Medina, A. et al. Unified metagenomic method for rapid detection of microorganisms in clinical samples. Communications Medicine 4, 135 (2024). 10.1038/s43856-024-00554-3

30 Campos-Madueno, E. I., Aldeia, C. & Endimiani, A. Nanopore R10.4 metagenomic detection of blaCTX-M/blaDHA antimicrobial resistance genes and their genetic environments in stool. Nature Communications 15, 7450 (2024). 10.1038/s41467-024-51929-y

31 Jing, D. et al. Metagenomic nanopore sequencing of ocular microbiome in patients with meibomian gland dysfunction. Frontiers in Medicine Volume 9 - 2022 (2022). 10.3389/fmed.2022.1045990

32 Nielsen, M. E. et al. Application of rapid Nanopore metagenomic cell-free DNA sequencing to diagnose bloodstream infections: a prospective observational study. Microbiology Spectrum 13, e03295–03224 (2025). doi:10.1128/spectrum.03295-24

33 Che, Y. et al. Mobile antibiotic resistome in wastewater treatment plants revealed by Nanopore metagenomic sequencing. Microbiome 7, 44 (2019). 10.1186/s40168-019-0663-0

34 Rautiainen, M. & Marschall, T. MBG: Minimizer-based sparse de Bruijn Graph construction. Bioinformatics 37, 2476–2478 (2021). 10.1093/bioinformatics/btab004

