## Supplementary Information for "Mind the Gaps: Validating species presence under sparse genomic recovery in clinical metagenomics"

### Supplementary Note 1. Statistical and Mathematical Rationale of MAVI

This note provides the formal foundations of the Metagenomic Alignment Validation Index (MAVI). The construction is a one-dimensional biological specialisation of the generator-level dispersion framework[1] and the Generator Identifiability Envelope (GIE)[2] developed for Boolean random closed sets, with genomic coordinates replacing Euclidean space and aligned reads providing the recovered spatial configuration. Supplementary Note 1.1 introduces the stochastic losses generated by sample preparation and sequencing. Supplementary Note 1.2 formalises the Monte Carlo full-support reference model and the compatibility coordinate derived from the observed dispersion $D$. Supplementary Note 1.3 introduces the operational $\alpha$-gate and then defines restricted-support mimicry under sparse recovery. Supplementary Note 1.4 explains why increasing genomic recovery progressively separates full- and restricted-support compatibility laws and produces a sigmoid-like transition with breadth. Supplementary Note 1.5 defines the $\beta_0$-controlled compatibility boundary $T(b)$, establishes its monotonicity over admissible breadth intervals and introduces the logistic approximation used for MAVI inversion in the main text.

#### 1.1 Stochastic characterisation of sample-preparation processes

A stochastic process is defined on a probability space $(\Omega, F, P)$, where $\Omega$ is the set of possible outcomes, $F$ a $\sigma$-algebra of measurable events, and $P$ a probability measure. A stochastic process is a family of random variables

$$X_t : \Omega \rightarrow S,\ t \in T\ ,$$

mapping each outcome $\omega \in \Omega$ to a state $X_t(\omega)$ in some state space $S$ ($S$ = (genomic fragment retained, genomic fragment lost)).

Sample-preparation steps such as cell lysis, DNA extraction, host-depletion and library preparation can each be represented as stochastic processes of this form. Their outcomes are random because the physical and biochemical mechanisms that retain or lose DNA fragments vary across molecules and across steps. The set of fragments that survive the full pipeline is therefore a random subset of the original genomic material. Sequencing reads constitute a realisation of this multi-stage stochastic process.

**Relevance to MAVI**

This formalism justifies treating the observed reads as the outcome of a random fragmentation process under $H_0$, providing the probabilistic foundation for the Monte Carlo null model used in MAVI.

#### 1.2 Monte Carlo assessment of compatibility with random fragmentation

Let $D$ denote the empirical dispersion of alignment blocks derived from mapped reads for a candidate genome of length $G$, given $x$ reads with empirical read-length distribution $r$. Under the null hypothesis $H_0$ (random retention of fragments) we simulate many synthetic datasets by placing $x$ reads uniformly across the genome, sampling lengths from $r$, and computing $D$ for each simulation. This yields statistics $D_1, D_2, \ldots, D_N$ with Monte Carlo mean and variance

$$\mu_{MC} = \frac{1}{N}\sum_{i}^{N} D_i$$
$$\sigma_{\mathrm{MC}}^2 = \frac{1}{N-1}\sum_{i=1}^{N}\left(D_i - \mu_{\mathrm{MC}}\right)^2.$$

We define the standardised deviation

$$Z = \frac{D_{obs} - \mu_{MC}}{\sigma_{MC}}$$

which quantifies how far $D_{\mathrm{obs}}$ departs from the Monte Carlo mean.

The standardised deviation $Z$ is mapped through $\Phi(Z)$, providing a continuous coordinate of compatibility with the candidate-specific full-support fragmentation model. The empirical Monte Carlo lower-tail probability

$$p_{\mathrm{MC}} = \frac{\#\{i : D_i \le D_{\mathrm{obs}}\} + 1}{N+1}$$

is additionally reported as a direct diagnostic of the null distribution.

**Remark** (Monte Carlo null-variance collapse). When $\sigma_{\mathrm{MC}} \mapsto 0$, $Z$ is undefined and $\Phi(Z)$ is set to 0.5 as a neutral placeholder. Variance collapse can occur in distinct limiting regimes, including near-empty and near-saturated recovery, and therefore carries no directional information about generator support by itself. The candidate is retained for MAVI inversion so that its support requirement is evaluated against the complete minimax compatibility boundary, rather than inferred from variance collapse alone. It is excluded from $H_{\mathrm{norm}}$ because the assigned $\Phi(Z)$ value is a computational placeholder rather than a data-derived compatibility value.

**Relevance to MAVI**
This section defines the continuous full-support compatibility coordinate $\Phi(Z_{\mathrm{obs}})$ used by the MAVI construction.

#### 1.3 Restricted-support mimicry under sparse recovery

In the operational implementation, a preliminary $\alpha$-gate removes candidates whose observed dispersion is already incompatible with random fragmentation across the full candidate genome. For a declared $\alpha$, candidates satisfying

$$\Phi\left(Z_{\mathrm{obs}}\right) < \alpha$$

are excluded before MAVI inversion. The $\alpha$-gate is therefore a computational screening step. It is not part of the definition of the minimax envelope, but reduces the computational burden by excluding taxa with low full-support compatibility before envelope inversion.

Passing this initial gate establishes only compatibility with full-support fragmentation at threshold $\alpha$. Under sparse recovery, restricted genomic support may produce the same or stronger compatibility, so passing the gate does not determine how much of the candidate genome must have been represented.

For a candidate support fraction $C_{\mathrm{f}}$ and compatibility threshold $t$, we quantify this ambiguity through the restricted-support mimicry probability

$$\beta\left(C_{\mathrm{f}}, t\right) = P_{H_1(C_{\mathrm{f}})}\left(\Phi(Z) \geq t\right).$$

With $H_1(C_{\mathrm{f}})$ being the restricted-support alternative. Thus, $\beta(C_{\mathrm{f}},t)$ measures how readily restricted genomic support can generate compatibility with full-genome fragmentation at or above threshold $t$. For an observed recovery, the relevant threshold is $t = \Phi(Z_{\mathrm{obs}})$.

Fixing a tolerated mimicry probability $\beta_0$ converts this family of probabilities into the compatibility boundary

$$T(C_{\mathrm{f}}) = \inf\left\{t \in [0,1] : \beta(C_{\mathrm{f}}, t) \leq \beta_0\right\}.$$

Thus, $T(C_\mathrm{f})$ is the full-support compatibility threshold corresponding to tolerated mimicry $\beta_0$ for restricted support $C_\mathrm{f}$. MAVI is obtained by inverting this boundary at the observed compatibility $\Phi(Z_\mathrm{obs})$, as developed below.

**Relevance to MAVI**

Under sparse recovery, restricted genomic support can reproduce compatibility with random full-genome fragmentation. The mimicry probability $\beta(C_\mathrm{f},t)$ quantifies how readily this can occur for a candidate support fraction $C_\mathrm{f}$ at compatibility threshold $t$. MAVI inverts this ambiguity to determine the minimum genomic support an alternative source would require to reproduce the observed dispersion evidence.

#### 1.4 Sigmoidal separation of full- and restricted-support fragmentation

Let the sampling distributions of $D$ under full genomic support $H_0$ and a restricted-support alternative $H_1(C_\mathrm{f})$ be denoted

$$\begin{aligned} D \mid H_0 &\sim f_0\left(d;x\right), \\ D \mid H_1(C_\mathrm{f}) &\sim f_1\left(d;x,C_\mathrm{f}\right), \end{aligned}$$

where $x$ indexes the amount of recovered information and induces the corresponding expected breadth $b(x)$. As $x$ increases, three behaviours occur:

1. the sampling variability of $D$ decreases under both support regimes;
2. the restricted- and full-support distributions separate relative to their variability;
3. the probability that restricted support reproduces strong full-support compatibility decreases smoothly.

The compatibility coordinate is

$$\begin{aligned} Y &= \Phi(Z), \\ Z &= \frac{D-\mu_0(x)}{\sigma_0(x)}, \end{aligned}$$

where $\mu_0(x)$ and $\sigma_0(x)$ are the full-support mean and standard deviation. Under a restricted-support alternative, dispersion is reduced relative to full genomic support, so $\mu_1(x) < \mu_0(x)$. As recovery increases through the informative regime, this separation becomes progressively larger relative to the sampling variability, shifting the restricted-support compatibility distribution towards lower values of $Y$.

Under a normal approximation,

$$D \mid H_1(C_\mathrm{f}) \approx N\left(\mu_1(x), \sigma_1^2(x)\right),$$

and therefore

$$Z \mid H_1(C_\mathrm{f}) \approx N\left(\delta(x), \rho^2(x)\right),$$

with

$$\delta(x) = \frac{\mu_1(x) - \mu_0(x)}{\sigma_0(x)}, \qquad \rho(x) = \frac{\sigma_1(x)}{\sigma_0(x)}.$$

For a fixed tolerated mimicry probability $\beta_0$, the corresponding upper compatibility boundary is therefore approximately

$$T(x \mid C_\mathrm{f}) \approx \Phi\left[\delta(x) + \rho(x)\Phi^{-1}\left(1 - \beta_0\right)\right].$$

At low recovery, full- and restricted-support fragmentation overlap strongly and the boundary remains high. Across the intermediate regime, increasing separation produces a rapid transition towards lower compatibility. Once the two regimes are well separated, further recovery produces progressively smaller changes. This gives the compatibility boundary its characteristic sigmoid-like trajectory. Because expected breadth $b(x)$ increases monotonically with recovered information, the same transition is expressed as a function of breadth.

**Relevance to MAVI**

This breadth-dependent separation motivates the logistic approximation of the $\beta_0$-controlled compatibility boundary $T(b \mid C_\mathrm{f})$ used to construct the MAVI envelope.

#### 1.5 Logistic approximation of the $\beta_0$-controlled compatibility boundary as a function of breadth

For a fixed tolerated mimicry probability $\beta_0$, let $Z_{1-\beta_0}(b)$ denote the upper (1- $\beta_0$)-quantile of the standardised dispersion distribution under the restricted-support alternative at breadth ($b$). The corresponding compatibility boundary is

$$T(b) = \Phi\left(Z_{1-\beta_0}(b)\right)$$

Thus, $T(b)$ is the compatibility value exceeded by a restricted-support alternative with probability at most $\beta_0$. The sigmoidal separation between the full- and restricted-support dispersion distributions established in Supplementary Note 1.4 implies that this boundary changes smoothly and sigmoidally with breadth.

#### *1.5.1 Monotonicity of the population compatibility boundary*

Fix $G$, $r$, $C_f$, and $s$, and let $Y_b = \Phi(Z_b)$ denote the population compatibility coordinate under the restricted-support alternative $H_1(C_f)$ at breadth $b$. For $t \in [0,1]$, define the corresponding mimicry probability $\beta(b,t) = \Pr_{H_1(C_f)}\{Y_b > t\}$.

For a declared tolerance $\beta_0 \in (0,1)$, the population compatibility boundary is

$$T(b) = \inf\{t \in [0,1] : \beta(b,t) \le \beta_0\}.$$

**Definition 1** (Admissible breadth interval). A breadth interval $\mathcal{B} \subseteq [0,1]$ is admissible if the restricted-support compatibility laws are stochastically ordered across breadth. That is, for every $b_1, b_2 \in \mathcal{B}$ with $b_1 \le b_2$, $Y_{b_2} \le_{\text{st}} Y_{b_1}$, or equivalently,

$$\beta(b_2,t) \le \beta(b_1,t) \qquad \text{for every } t \in [0,1].$$

This condition states that increasing breadth cannot increase the probability that a restricted-support generator attains any declared level of full-support compatibility.

**Proposition 1** (Monotonicity in breadth). On every admissible breadth interval $\mathcal{B}$, the map $b \mapsto T(b)$ is non-increasing.

**Proof.** For each $b \in \mathcal{B}$, define $E(b) = \{t \in [0,1] : \beta(b,t) \le \beta_0\}$. Let $b_1 \le b_2$. By stochastic ordering,

$$\beta(b_2,t) \le \beta(b_1,t) \qquad \text{for every } t.$$

Therefore, every threshold satisfying the mimicry criterion at $b_1$ also satisfies it at $b_2$, so $E(b_1) \subseteq E(b_2)$. Consequently,

$$T(b_2) = \inf E(b_2) \le \inf E(b_1) = T(b_1).$$

Hence $T(b)$ is non-increasing on $\mathcal{B}$.

The breadth-ordering condition is the distributional form of the increasing separation developed in Supplementary Note 1.4. The breadth-ordering condition directly orders the complete compatibility law and therefore guarantees monotonicity for any fixed $\beta_0$. The result applies on the non-degenerate breadth interval over which increasing genome representation progressively reduces the mimicry capacity of restricted-support alternatives.

Because the exact mapping from $b$ to $T(b)$ is not analytically tractable, we approximate it using the logistic function

$$T(b) = \frac{1}{1+e^{-k(b-b_0)}}, \qquad k < 0.$$

Here $b_0$ is the midpoint of the compatibility transition and $k$ controls the steepness of the transition. Together, these parameters provide a compact representation of the expected sigmoidal behaviour of the $\beta_0$-controlled compatibility boundary. This boundary is the one-dimensional genomic form of the support-fraction compatibility boundary underlying the Generator Identifiability Envelope[2].

**Relevance to MAVI**
This logistic $T(b)$ represents the $\beta_0$-controlled compatibility boundary used to determine the minimum restricted genomic support capable of reproducing a candidate's observed breadth–$\Phi(Z)$ pair.

### Supplementary Note 2 Choice of the merging threshold *s*

This analysis specifies the admissible dispersion scales $s$ that correspond to the non-degenerate window required by the stabilisation and entropy-collapse conditions of the generator-level dispersion diagnostics[1,2].

Let a genome of length $G$ contain $x$ mapped reads with empirical read-length distribution $r$. For a candidate genome, the dispersion statistic $D$ depends on the number of gaps between consecutive reads that exceed the threshold $s$. Formally, under the standard Poisson approximation with intensity $x/G$, the inter-read start spacings are approximately exponential with mean

$$\mu = \frac{G}{x}$$

Thus, the probability that a gap is shorter than $s$ is

$$p(s) = 1 - e^{-s/\mu}$$

Two limiting regimes follow directly:

1. If $s \ll \mu$, then $p(s) \approx s/\mu$ and almost every gap exceeds $s$.
   The expected dispersion is approximately $D \approx x$, making clustered and non-clustered patterns indistinguishable.
2. If $s \gg \mu$, then $p(s) \approx 1$ and almost all gaps are merged.
   The dispersion collapses to $D \approx 1$, eliminating discriminatory power.

Hence informative behaviour of $D$ occurs only when $s$ is of order $\mu$ or, more generally, when $s$ lies strictly between the scales of clustered and non-clustered gaps. When $s$ is smaller than $\mu$, the large genome-wide gaps expected under $H_0$ remain distinct, preventing over-merging and preserving the characteristic block structure that $D$ is designed to detect. However, $s$ must not be taken arbitrarily small: if $s$ is much smaller than the minimum gap lengths generated by read placement noise, even closely spaced reads are treated as separate blocks, driving $D$ artificially towards $x$ and erasing any contrast between clustered and uniform patterns. If $s$ approaches or exceeds $\mu$, most $H_0$ gaps collapse and the dispersion statistic loses discriminatory power. Thus, $D$ is informative only when $s$ lies between these two limits: small enough to avoid collapsing the large $H_0$ gaps, yet large enough to avoid fragmenting trivial positional fluctuations. Maintaining $s$ in this intermediate regime, with $s < \mu$ but above the lower fluctuation scale, ensures that the dispersion functional operates at its stabilisation scale and preserves its discriminatory structure.

In clinical metagenomics, genome sizes $G$ are typically in the order of $10^6$ bp and the number of mapped reads per candidate genome $x$ is in the order of $10^2$–$10^3$. Under random genome-wide fragmentation, the expected spacing between read start positions is $\mu = G/x$, which therefore lies in the order of $10^3$–$10^4$ bp. In our dataset, most genomes fall between 1 and 5 million base pairs and only a few hundred reads map to each genome. Taking a conservative value of $x = 250$ gives $\mu \approx 4000$–$20000$ bp across this range of $G$. This defines the upper scale at which gaps may still be regarded as consistent with random full-genome coverage, and therefore provides an upper constraint for the merging threshold $s$.

A consistent conclusion follows directly from coverage. The maximum possible coverage implied by $x$ reads of length $r$ on a genome of length $G$ is $c_{\max} = xr/G$, because overlaps between reads make the realised coverage strictly smaller. Rearranging gives $\mu = G/x = r/c_{\max}$, showing that under $H_0$ the expected inter-read gap is bounded below by the read length divided by the maximum achievable coverage. With read lengths in the range 600–6000 bp and actual coverage well below 1 in clinical metagenomic samples, $\mu$ necessarily lies in the $10^3$–$10^4$ bp range. Even adopting an optimistic value of $c = 0.5$, which exceeds typical clinical metagenomic coverage, one still obtains $\mu = r/c_{\max} \approx 2r$, keeping $\mu$ in the same $10^3$–$10^4$ bp regime. Moreover, standard long-read metagenomic practice excludes reads shorter than roughly 500 bp; this imposes a hard lower bound of about 1000 bp on $\mu$. These constraints mean that $\mu$ is unlikely to fall below ~1 kb for any realistic combination of $G$, $x$ or $r$. Hence $s = 1000$ bp satisfies $\mu > s$ for all relevant cases compatible with $H_0$.

The simulations confirm the behaviour predicted in the theoretical analysis. Reducing $s$ (e.g. to 500 bp) increases the influence of small positional fluctuations between reads, which produces slightly larger scatter in $\Phi(Z)$ at low breadth. Increasing $s$ (e.g. to 2000 bp) merges reads more aggressively and therefore shifts $\Phi(Z)$ towards the upper boundary at high breadth. However, in both cases the effect is purely local and does not alter the logistic boundary $T(b)$, which encapsulates the decision-relevant behaviour of the test (Fig. S2.1).

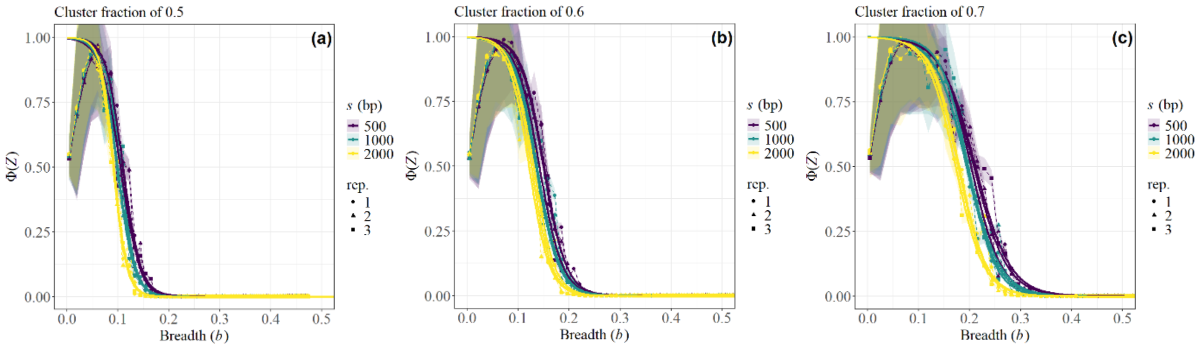


Fig. S2.1 | Panels a–c show $\Phi(Z)$ as a function of breadth ($b$) for cluster fractions $C_f = 0.5$, 0.6 and 0.7 under three merging thresholds ($s = 500$, 1000 and 2000 bp). Points represent empirical (1- $\beta_0$)-quantile boundary estimates at the simulated read-count grid points; point shapes distinguish independent simulation runs, shaded ribbons show variability across runs, and solid curves show the fitted logistic boundaries $T(b)$.

Crucially, the logistic boundary is the object used for inference. It represents the $\Phi(Z)$ threshold above which a candidate cannot plausibly arise from clustered alternatives under $\beta_0 = 0.001$. Because $T(b)$ is monotonic and smooth, small fluctuations in $\Phi(Z)$ induced by different merging thresholds do not propagate into meaningful differences in MAVI values (Fig. S2.2). In other words, while the raw $\Phi(Z)$ curves show the expected stochastic variability, their corresponding logistic boundaries sit almost exactly on the same trajectory.

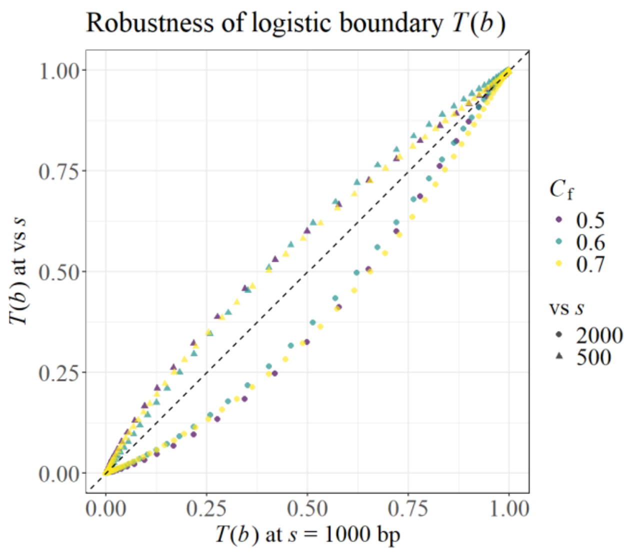


Fig. S2.2 | Comparison of logistic boundaries across merging thresholds. For each $b$, $G$, $r$ and $C_f$ value, the fitted values $T(b; s = 500)$ and $T(b; s = 2000)$ are plotted against $T(b; s = 1000)$. Colours denote cluster fractions $C_f$, and point shapes indicate the $s$-pair being compared. The dashed line marks the 1:1 relationship.

This behaviour has a straightforward interpretation. For cases lying very close to $T(b)$, changing $s$ may shift $\Phi(Z)$ slightly above or below the boundary, but such cases are not categorically decided in the first place. MAVI yields a continuous support-fraction bound rather than a hard classification, so small movements around the transition region do not represent a real change in the strength of evidence. Quantitatively, varying $s$ between 500 and 2000 bp shifts the logistic midpoint $b_0$ by at most ~0.025 in breadth (typically ~0.01), and changes in logistic steepness $k$ are modest (median 3% for $s = 500$ and ~16% for $s = 2000$). All pseudo-$R^2$ values remain above 0.98, confirming that the sigmoidal form is preserved. Across all realistic combinations of $G$, $x$ and $r$, and across $C_f = 0.5$–$0.7$, the ordering of candidates relative to $T(b)$ is therefore unchanged, and the practical decision power of the method remains stable.

Taken together, these scales imply that $s$ must lie well below the smallest $\mu$ that can arise under $H_0$, yet remain large enough not to fragment minor fluctuations in read placement[2]. This confines $s$ to a narrow window in the low-kilobase range: the lower bound is dictated by the minimum feasible $\mu$ of roughly 1 kb, whereas the upper bound is set by genome-wide random gaps of several kilobases (typically 4–20 kb). Within this interval, choosing $s = 1000$ bp places the merging threshold exactly where it is most informative. It is low enough to preserve the large-scale gap structure expected under $H_0$, ensuring that the block patterns detected by $D$ are not lost, yet high enough to avoid treating tightly spaced reads as separate blocks. The simulation results in Figs. S2.1–S2.2 confirm that $T(b)$ remains stable throughout this range, reinforcing that $s$ = 1000 bp is both physically justified and empirically robust.

**Relevance to MAVI**

The analysis shows that $s$ = 1000 bp aligns with the natural spacing scales expected under $H_0$. Clinical metagenomic datasets typically contain a few hundred to thousand reads dispersed over genomes of megabase size, yielding inter-read spacings of several kilobases. A threshold of 1000 bp preserves these large, genome-scale gaps that characterise random whole-genome coverage, while smoothing over minor positional fluctuations that do not correspond to meaningful fragmentation. In this position, $s$ stabilises the dispersion statistic $D$ and ensures reliable Monte Carlo behaviour across the realistic ranges of $G$, $x$ and $r$ encountered in clinical metagenomic applications.

### Supplementary Note 3. Logistic modelling of the $\beta_0$-controlled compatibility boundary

This note implements the minimax identifiability-envelope construction of the Generator Identifiability Envelope[2] within the genomic setting used by MAVI. For each combination of genome size $G$, empirical read-length distribution $r$, and support fraction $C_f$, we estimate the $\beta_0$-controlled compatibility boundary

$$T(b(x) \mid C_f) = Q_{1-\beta_0}\left[\Phi(Z_x) \mid H_1(C_f)\right]$$

where $Q_{1-\beta_0}$ denotes the upper (1-$\beta_0$)-quantile of the compatibility distribution generated by the restricted-support alternative. Equivalently,

$$\Pr_{H_1(C_f)}\left\{\Phi(Z_x) > T\big(b(x) \mid C_f\big)\right\} \le \beta_0.$$

Throughout the analyses, $\beta_0$=0.001, and the boundary is abbreviated as $T(b \mid C_f)$. Supplementary Note 3.1 estimates this boundary using order statistics from restricted-support Monte Carlo simulations. Supplementary Note 3.2 maps the resulting boundary values to empirical breadth and identifies the monotonic modelling region. Supplementary Note 3.3 fits the corresponding logistic approximation by maximum quasi-likelihood. Throughout, $x$ denotes the number of reads and $N$ the number of Monte Carlo replicates for a given $x$.

#### 3.1 Empirical $\beta_0$-controlled compatibility boundary under clustered alternatives

Fix $G$, $r$ and a cluster fraction $C_f \in (0, 1]$. Consider a grid of read counts

$$x \in \{x_1, x_2, \ldots, x_m\}$$

with $x_1 < x_2 < \ldots < x_m$. For each $x$ we perform two sets of 100,000 Monte Carlo simulations:

1. Null simulations (random fragmentation): reads are placed uniformly at random across the full genome [0, $G$] with read lengths drawn from $r$. For each replicate $j = 1, \ldots, N$ we compute the dispersion statistic $D_j^0(x)$. This yields a null Monte Carlo sample $\{D_j^0(x)\}_{\{j=1\}^N}$ with empirical mean $\mu_{MC}(x)$ and standard deviation $\sigma_{MC}(x)$.
2. Clustered simulations: reads are placed uniformly at random within a contiguous region of length $C_f G$, again with read lengths drawn from $r$ and using the same dispersion definition. For each replicate $j = 1, \ldots, N$ we obtain a clustered dispersion value $D_j^1(x)$ and an empirical breadth $B_j^1(x)$ measured as the proportion of bases in [0, $G$] covered by at least one read under that replicate.

For each clustered replicate we define the corresponding $Z$-score

$$Z_j(x) = \frac{D_j^1(x) - \mu_{MC}(x)}{\sigma_{MC}(x)}$$

Let $Z_1(x) \le Z_2(x) \le \ldots \le Z_N(x)$ denote the ordered $Z_j(x)$ values. For a fixed mimicry tolerance $\beta_0$ we define the $(1 - \beta_0)$-quantile of the clustered $Z$ distribution via the order statistic

$$Z_{1-\beta_0}(x) = Z_{\lceil (1-\beta_0)N \rceil}(x).$$

With $\beta_0 = 0.001$ we obtain $Z_{0.999}(x)$. The associated empirical $\beta_0$-controlled compatibility boundary is

$$\hat{T}(x \mid C_f) = \Phi(Z_{0.999}(x)),$$

where $\Phi$ denotes the standard normal cumulative distribution function.

The contiguous cluster is a computational representation of restricted genomic support, not an assumption that shared loci are physically adjacent. Partitioning the same support into $v_s$ components separated at scale ($s$) can increase the dispersion statistic by at most $v_s$-1, and may therefore produce a high $\Phi(Z)$ when only a few reads are observed. However, $\Phi(Z)$ is only the compatibility coordinate; certification requires inversion against the $\beta_0$=0.001-controlled restricted-support boundary. In the sparse regime, where a small number of separated reads can most strongly inflate dispersion, this boundary lies close to one and such configurations remain uncertified. For fragmentation to continue mimicking random genome-wide recovery as breadth increases, $v_s$ must grow on the same scale as the observed dispersion. The retained support is then itself broadly distributed across genomic coordinates and is no longer a narrow cluster masquerading as species-level representation. Fragmentation therefore either fails the MAVI boundary or defines a genuine identifiability limit that the boundary correctly preserves. This is the hidden-support logic of the Generator Identifiability Envelope: compatibility with random fragmentation is insufficient unless it cannot be reproduced, at the declared mimicry tolerance, by an admissible restricted support, including a fragmented support whenever fragmentation is the least-favourable configuration[2].

**Remark** (physical scope of the fragmented-support argument). By Proposition 3 of the GIE framework[2], the maximal geometry-free support class may be restricted when additional structural information constrains the hidden supports admissible under the observation regime. In MAVI, finite genome length, read count, read length and separation scale provide exactly such information. Accordingly, the treatment of cases with $v_s>1$ above is relative to the admissible long-read observation regime, rather than to an unrestricted class of abstract support geometries. Let $\ell_{\min} = \min \operatorname{supp}(r) > 0$ denote the minimum read length in the empirical read-length distribution. For finite genome length $G$, finite read count $x$, and fixed separation scale $s>0$, every represented component must contain at least one read of length at least $\ell_{\min}$, while components counted separately must be separated by at least $s$. Consequently,

$$v_s \leq \min\left\{x, \left\lfloor \frac{G+s}{\ell_{\min}+s} \right\rfloor\right\} < \infty.$$

For the long-read regime considered here, with $G$ typically between 1 and 5 $10^6$ bp, $s=1000$ bp, and $\ell_{\min} \approx 500$ bp, the geometric packing bound lies between approximately 667 and 3334 separated components. In practice, however, each represented component must contain at least one mapped read and remain separated from the others at scale $s$. The number of components that can influence the observed dispersion is therefore bounded by the read count $x$, which is usually only a few hundred. Approaching this bound would require nearly one read per separated component, making such a configuration intrinsically fragile: merging even a few components directly reduces dispersion and weakens its ability to mimic full-genome fragmentation. The read-count constraint is therefore reached long before the geometric packing limit.

Supports requiring arbitrarily many infinitesimal disconnected components or read lengths tending to zero are therefore not admissible competing generators under this observation regime: neither the required fragments nor the number of reads needed to represent them exist. The argument above is uniform over the physically admissible long-read class; it is not asserted for an enlarged abstract class in which $\ell_{\min} \downarrow 0$ or infinitely fragmented supports are permitted.

Because breadth is the quantity of biological interest, we map each read count *x* to an empirical breadth by averaging the observed clustered breadth across replicas:

$$b(x) = \frac{1}{N}\sum_{j=1}^{N} B_j^1(x)$$

Because breadth is a saturation statistic, its Monte Carlo variability is intrinsically small: once reads begin to overlap within the clustered region, the realised breadth concentrates tightly around its expectation. Consequently, the $x \mapsto b(x)$ introduces negligible noise and does not affect the stability of the $\Phi(Z)$–breadth calibration.

This defines a set of points

$$\left(b_i, \hat{T}_i\right) = \left(b(x_i), \hat{T}(x_i)\right), i = 1, \ldots, m$$

which form the empirical breadth– compatibility boundary $\hat{T}(b)$ under the clustered model with parameters ($G$, $r$, $C_f$).

**Relevance to MAVI**

This construction links restricted-support mimicry to the biologically interpretable quantity of breadth. For each degree of clustering $C_f$, it shows how the compatibility index $\Phi(Z)$ evolves as genome coverage increases. In this way, it quantifies how strongly clustered alignments would continue to resemble the full-support model at each empirical breadth value.

Using empirical breadth $b(x)$ incorporates the effects of read overlap and coverage pile-up directly into the analysis. This provides a biologically grounded relationship between genome representation and the $\Phi(Z)$ values that MAVI uses to determine how much restricted genomic support is required to reproduce the observed compatibility.

#### 3.2 Identification of the monotonic modelling region

By Proposition 1 in Supplementary Notes 1.5, the population compatibility boundary $T(b)$ is non-increasing on every admissible breadth interval. Its finite Monte Carlo estimator $\hat{T}(b)$, however, may contain local violations of this order because the upper quantiles are estimated from finitely many simulations. Moreover, near breadth saturation the admissibility condition itself may cease to hold as the full-support and restricted-support dispersion laws enter a degenerate regime. We therefore identify the empirical interval over which the population breadth ordering is stably represented before fitting the logistic model.

Let $b_1 < b_2 < \cdots < b_m$ denote the empirical breadth values obtained in Supplementary Note 3.1, with corresponding boundary values $\hat{T}_i = \hat{T}(b_i)$. Define the first differences

$$\Delta_i \hat{T} = \hat{T}_{i+1} - \hat{T}_i, \qquad i = 1, \ldots, m-1.$$

The lower boundary of the modelling region is defined by the first discrete local maximum of the empirical boundary. Specifically,

$$i_L = \min\left\{ i \in \{2, \ldots, m-1\} : \Delta_{i-1}\hat{T} > 0 \text{ and } \Delta_i \hat{T} < 0 \right\},$$

and $b_L = b_{i_L}$.

This identifies the first change from positive to negative slope and therefore the beginning of the decreasing compatibility transition. If no such sign change is detected, $b_L$ is set to the smallest empirical breadth. Breadth values below $b_L$ are excluded from the logistic fit.

The upper extent of the simulation grid is determined through the expected sequencing depth within the restricted support,

$$\lambda = \frac{xr}{C_f G}.$$

To ensure that the simulated breadth values span the compatibility transition, we set $\lambda_{\text{target}} = 3$, giving the maximum simulated read count

$$x_{\max} = \left\lfloor \frac{C_f G}{r} \lambda_{\text{target}} \right\rfloor.$$

This allows the restricted region to approach breadth saturation while avoiding unnecessarily large read-count grids.

An additional upper-tail diagnostic is applied to the empirical boundary values satisfying $b_i \geq b_L$. Let $\tilde{T}_i$ denote the isotonic reference sequence and define the residuals $e_i = \hat{T}_i - \tilde{T}_i$.

Two complementary criteria are used to detect an early departure from the stable trajectory.

First, let $\bar{e}$ and $s_e$ denote the mean and standard deviation of the residuals. A sustained departure is identified when nine consecutive residuals satisfy $e_i > \bar{e} + 8s_e$. The breadth at the beginning of the first such sequence defines the candidate upper cutoff $b_A$. If no sustained departure is detected, $b_A = \infty$.

Second, define the successive residual changes $\delta_i = e_{i+1} - e_i$ and $m_\delta = \text{median}_i |\delta_i|$. The first residual change satisfying $\delta_i > 5m_\delta$ defines the candidate cutoff $b_B$. If no such change is detected, $b_B = \infty$.

Let $b_{\max}$ denote the largest simulated breadth. The preliminary upper boundary is

$$b_U^{(0)} = \min\{b_A, b_B, 0.75, b_{\max}\}.$$

The residual criteria act as safeguards against an early onset of upper-tail instability, while the fixed limit $b = 0.75$ prevents the fit from extending into the near-saturation region. When the simulated grid does not reach 0.75, the maximum available breadth provides the effective upper limit.

To prevent a local fluctuation from producing an interval too narrow for stable estimation, a minimum modelling width of 0.5 breadth units is imposed:

$$b_U = \begin{cases} \min\{b_L + 0.5, b_{\max}\}, & b_U^{(0)} - b_L < 0.5, \\ b_U^{(0)}, & b_U^{(0)} - b_L \geq 0.5. \end{cases}$$

The final index set used for logistic fitting is $\mathcal{F} = \{i : b_L \le b_i \le b_U\}$. If $|\mathcal{F}| < 3$, the retained data contain too few points for stable estimation and the logistic parameters are not fitted for that combination of $G$, $r$, and $C_\mathrm{f}$.

**Relevance to MAVI**
This procedure isolates the portion of the empirical compatibility boundary in which the breadth-dependent separation of full-support and restricted-support fragmentation is stably represented. The lower cutoff removes the sparse region preceding the directional compatibility transition, while the upper safeguards prevent saturation-related instability from determining the logistic fit. The retained interval therefore captures the central sigmoidal region used to quantify how rapidly restricted-support explanations become implausible in MAVI.

**3.3 Logistic model for the compatibility boundary**

For each filtered index $i \in F$ we define $p_i = \hat{T}(b_i)$. We approximate the relationship between breadth and the compatibility boundary by a logistic function $T(b, k, b_0)$ (see Supplementary Notes 1.5), where $b_0 \in (0, 1)$ is the inflection point (breadth at which the compatibility boundary equals 0.5) and $k < 0$ controls the steepness of the transition.

To estimate $(b_0, k)$ from the empirical pairs $(b_i, p_i)$ we fit a logistic regression model

$$\operatorname{logit}(p_i) = \gamma_0 + \gamma_1 b_i + \varepsilon_i,\ i \in F$$

where $\operatorname{logit}(p) = \ln(p / (1 - p))$ and $\varepsilon_i$ is a mean-zero error term with quasi-binomial variance and dispersion parameter $\varphi$. Estimation proceeds by maximum quasi-likelihood.

The logistic parameters $(b_0, k)$ are related to the regression coefficients $(\gamma_0, \gamma_1)$ by

$$\hat{k} = \hat{\gamma}_1, \qquad \hat{b}_0 = -\frac{\hat{\gamma}_0}{\hat{\gamma}_1}.$$

Substituting these expressions into the logistic form yields the fitted compatibility boundary $\hat{T}(b, \hat{k}, \hat{b}_0)$.

**Relevance to MAVI**
The logistic model provides a compact, interpretable summary of how the compatibility threshold attainable by clustered explanations decreases with breadth for a given level of clustering $C_\mathrm{f}$. The parameter $\hat{b}_0$ locates the breadth at which the evidence crosses the midpoint of the compatibility transition, and $\hat{k}$ quantifies how sharply the transition occurs. Together

they encode how quickly clustered alternatives become less plausible as more of the genome is represented.

#### 3.4 Deviance and pseudo-$R^2$

To assess the adequacy of the logistic approximation we compute a deviance-based pseudo-$R^2$. Let $\hat{p}_i = \hat{T}(b_i)$ denote the fitted boundary values from the logistic model at breadth $b_i$. The logistic deviance is defined as

$$Dev = 2\sum_{i\in F}\left[p_i \ln\left(\frac{p_i}{\hat{p}_i}\right) + (1-p_i)\ln\left(\frac{1-p_i}{1-\hat{p}_i}\right)\right].$$

The null deviance $Dev_0$ is obtained by replacing $\hat{p}_i$ with the empirical mean

$$\bar{p} = \frac{1}{|F|}\sum_{i\in F} p_i ,$$

for all $i \in F$. The deviance-based pseudo-$R^2$ is then

$$R^2 = 1 - \frac{Dev}{Dev_0}.$$

Values of $R^2$ close to 1 indicate that the filtered empirical breadth–compatibility boundary is well described by the logistic form, consistent with the expected sigmoidal behaviour of the $\beta_0$-controlled compatibility boundary. Values near 0 indicate that the curve is nearly flat or highly irregular, and the corresponding logistic parameters are less informative.

**Relevance to MAVI**

The deviance-based pseudo-$R^2$ quantifies how well the logistic boundary captures the simulated compatibility-boundary behaviour for each combination of $G$, $r$ and $C_f$. High $R^2$ values provide reassurance that the compatibility boundaries used later in MAVI are not artefacts of the chosen parametric form but accurately reflect the underlying Monte Carlo behaviour of clustered genomes.

### Supplementary Note 4 Distinct roles and selection of $\alpha$ and $\beta_0$

The parameters $\alpha$ and $\beta_0$ control distinct stages of MAVI.

The first stage evaluates compatibility with the full-support null hypothesis $H_0$: random fragmentation across the full candidate genome. A candidate is rejected at this stage when $D_{obs} < D_\alpha$ or equivalently $\Phi_{obs} < \alpha$.

Thus, $\alpha$ controls the probability of rejecting the full-support fragmentation model when that model is true. Candidates satisfying $\Phi_{obs} \geq \alpha$ are carried forward, but passing this gate does not itself certify genome-wide presence.

The second stage evaluates the restricted-support alternatives $H_1(C_f)$: random fragmentation within a genomic support fraction $C_f$. For each breadth ($b$) and support fraction $C_f$, the compatibility boundary is defined by

$$T(b \mid C_f) = \inf\left\{ t \in [0,1] : \Pr_{H_1(C_f)}\left(\Phi(Z) > t \mid b\right) \leq \beta_0 \right\}.$$

Accordingly, $\beta_0$ is the tolerated upper bound on the probability that a restricted-support alternative exceeds the compatibility boundary. It does not define the initial $\alpha$-gate and is not the Type II error of that gate.

We set $\alpha$=0.001 as a conservative computational gate that removes candidates showing decisive evidence against random full-genome fragmentation before the more intensive MAVI inversion. This gate is operational rather than constitutive: the MAVI boundary remains well defined without it, but evaluating candidates that already fail the null-compatibility test adds computation without altering the inferential conclusion. Independently, we set $\beta_0$=0.001 so that certification requires a compatibility value that restricted-support alternatives reproduce with probability no greater than 0.1%. The equal numerical values are therefore a deliberately stringent choice at two separate stages and do not imply any mathematical or inferential coupling between $\alpha$ and $\beta_0$.

As with any decision framework, the numerical choices of $\alpha$ and $\beta_0$ are situational and may vary across clinical or operational contexts. Changing either value alters the strictness of the corresponding stage but does not affect the underlying logic of the approach: $\alpha$ controls early inclusion into MAVI, while $\beta_0$ governs the restricted-support mimicry tolerance used in MAVI inversion.

To illustrate the effect of $\beta_0$ on the logistic boundary, Fig. S4.1 shows the fitted $\Phi(Z)$–breadth curves for $\beta_0$ = 0.001, 0.01, 0.05 and 0.1 across three representative cluster-fraction values.

The curves differ slightly in their midpoint but remain similar in overall shape, especially in the region where decisions are made.

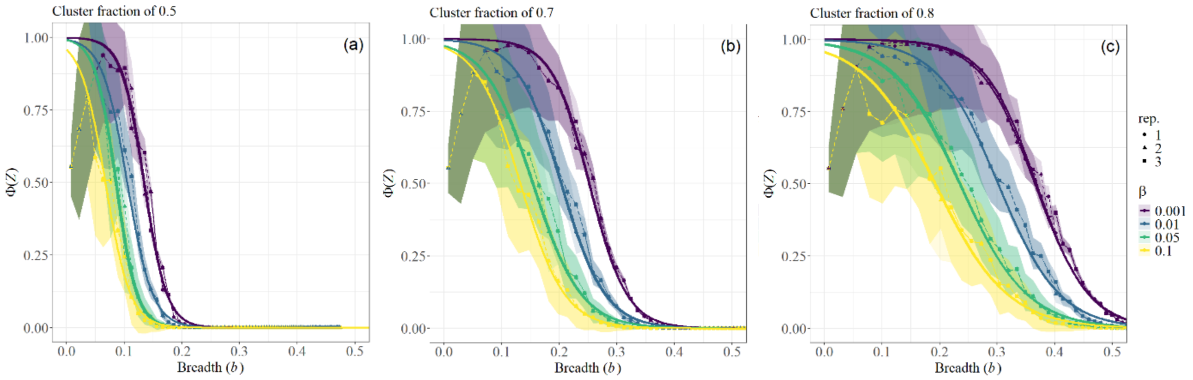


Fig. S4.1 | $\Phi(Z)$–breadth curves for $\beta_0 = 0.001, 0.01, 0.05$ and $0.1$ across cluster fractions $C_f = 0.5, 0.7$ and $0.8$. Each panel shows $\Phi(Z)$ as a function of breadth for the four $\beta_0$ values. Each point represents the empirical (1-$\beta_0$)-quantile compatibility boundary estimated from 100,000 clustered Monte Carlo replicates; dashed lines show replicate-specific trends; solid curves show the fitted logistic boundaries $T(b)$ for each $\beta_0$; shaded regions denote variability across repeats.

To examine this more directly, Fig. S4.2 compares the logistic boundaries $T(b)$ obtained with $\beta_0 = 0.001, 0.01, 0.05$ and $0.1$. Values of $T(b)$ computed with $\beta_0 = 0.01$ and $0.1$ follow smooth, systematic curves when plotted against $T(b)$ at $\beta_0 = 0.001$. Although the absolute threshold shifts with $\beta_0$, the boundary behaviour across $C_f$ remains consistent, confirming that the compatibility boundary is structurally stable across this range.

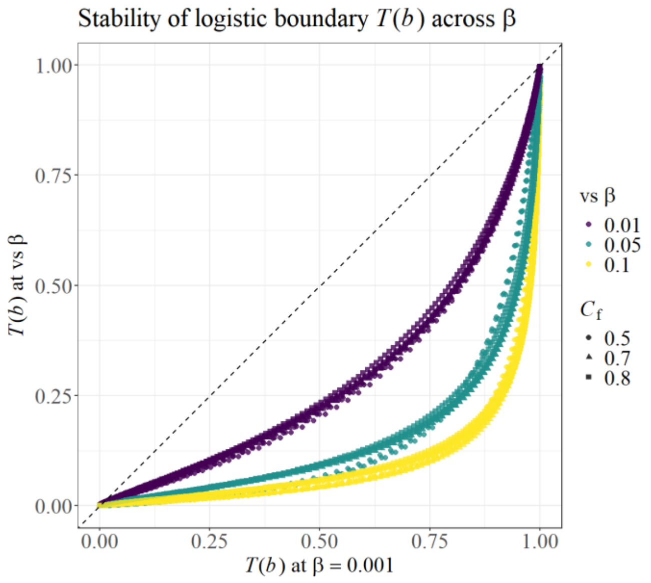


Fig. S4.2 | Comparison of logistic boundaries $T(b)$ obtained at $\beta_0 = 0.001$, $\beta_0 = 0.01$ and $\beta_0 = 0.1$. Each point represents $T(b)$ evaluated for a specific $(G, r, C_f, b)$ combination. Values obtained with $\beta_0 = 0.01$ or $\beta_0 = 0.1$ are plotted against those obtained at $\beta_0 = 0.001$. Colours denote the $\beta_0$ comparison; shapes denote the cluster fraction $C_f$. Points lie close to the 1:1 line, indicating that the logistic boundary is stable across $\beta_0$ values and that the ordering of candidates with respect to $T(b)$ is preserved.

Across all cluster-fraction values, the simulations show that changing $\beta_0$ between 0.001 and 0.1 systematically shifts the logistic boundary $T(b)$: smaller $\beta_0$ raises the curve and imposes a more stringent mimicry criterion, whereas larger $\beta_0$ lowers it and permits a greater probability of restricted-support mimicry. This is visible in Figs. S4.1–S4.2 as a controlled displacement of the transition region. Because MAVI is a continuous measure rather than a hard classifier, altering $\beta_0$ also changes the absolute MAVI values: increasing $\beta_0$ raises MAVI for borderline configurations, whereas decreasing $\beta_0$ lowers it. This behaviour is expected: $\beta_0$ specifies the tolerated upper bound on restricted-support mimicry, and the MAVI score responds smoothly rather than abruptly. Crucially, the relative ordering of candidates is preserved and the sigmoidal form of $T(b)$ remains intact. Thus, varying $\beta_0$ changes the strictness of the compatibility boundary in a predictable way, and choosing $\beta_0 = 0.001$ provides a deliberately conservative setting that strongly limits tolerated mimicry without introducing unstable or erratic behaviour.

The two controls therefore act on different inferential objects: $\alpha$ governs rejection of the full-support null at the initial dispersion gate, whereas $\beta_0$ governs the worst-case compatibility attainable by restricted-support alternatives during MAVI inversion.

**Relevance to MAVI**
In the analyses presented here, the initial compatibility threshold and the restricted-support mimicry tolerance were fixed independently at $\alpha$=0.001 and $\beta_0$=0.001. The former defines the preliminary gate for compatibility with the full-support fragmentation model; the latter limits the probability that a restricted-support alternative exceeds the compatibility boundary used for MAVI inference. Their equal numerical values impose stringent criteria at both stages but have distinct inferential roles.

### Supplementary Note 5 Parametric expressions for $b_0(G, r, C_f)$ and $k(G, r, C_f)$

The logistic curve used to relate breadth b to the corresponding $\beta_0$-controlled compatibility boundary (see Supplementary Note 1.5) contains the variables $b_0$ and $k$. To apply this curve analytically, $b_0$ and $k$ must be expressed as functions of genome size $G$, read length $r$ and cluster fraction $C_f$. Simulation results showed that both parameters vary smoothly across the parameter domain and can be accurately represented by simple linear relations on transformed scales.

The position parameter $b_0$ remains strictly positive, and its variation with $G$ and $r$ is approximately multiplicative. A log transformation therefore provides an adequate linearisation. Empirical inspection also showed that the effect of $C_f$ is non-linear but well captured by including $C_f$ and $C_f^2$. The fitted expression for $b_0(G, r, C_f)$ is

$$\ln(b_0) = a_0 + a_1 \log_{10}(G) + a_2 \log_{10}(r) + a_3 C_f + a_4 C_f^2 + a_5 \log_{10}(G) C_f + a_6 \log_{10}(r) C_f$$

with $b_0$ obtained as $e^{\ln(b_0)}$.

The model explains 99.7% of the variance in ln($b_0$) in the training simulations, with residual error approximately 0.06 on the log scale (Table S5.1).

Table S5.1 Coefficients for the $b_0$ model (ln($b_0$) scale)

| Coefficient | Estimate | StdError |
|---|---|---|
| Intercept | -1.213937 | 0.099304 |
| $\log_{10}(G)$ | -1.165382 | 0.008638 |
| $\log_{10}(r)$ | 1.530590 | 0.026348 |
| $C_f$ | 0.088429 | 0.166087 |
| $C_f^2$ | 2.525292 | 0.049761 |
| $\log_{10}(G)$:$C_f$ | 0.334854 | 0.013657 |
| $\log_{10}(r)$:$C_f$ | -0.472190 | 0.041659 |
| *Model Residual SE*: 0.05805 | | |
| $R^2$: 0.9965, adj. $R^2$: 0.9965 | | |

Observed and predicted values (Fig. S5.1) fall tightly along the 1:1 line, while the diagnostic plots (Fig. S5.2) show adequate model behaviour across the central fitted range, with departure from normality confined mainly to the upper residual tail.

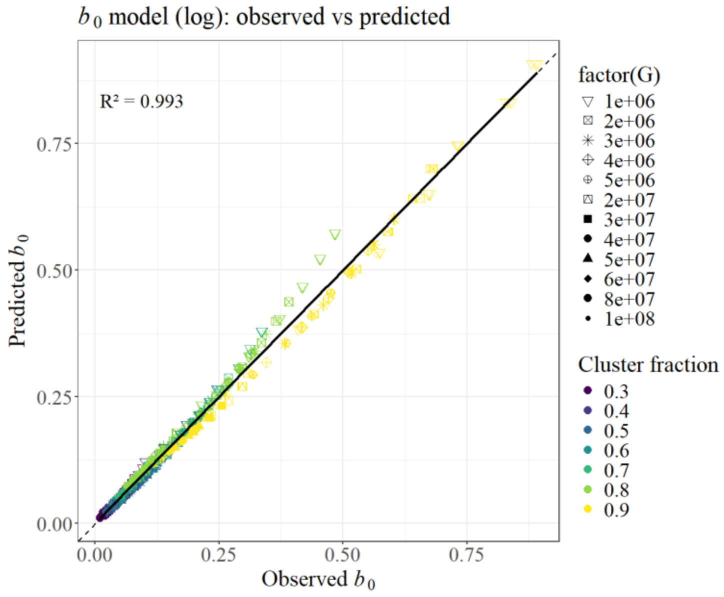

Fig. S5.1 | Observed versus predicted values of $b_0$. Observed $b_0$ from the simulation set used for model fitting (calibration run) plotted against predicted $b_0$ obtained from the parametric expression in Supplementary Note 5. Colours indicate cluster fraction and shapes indicate genome size. The dashed line denotes the 1:1 relation.

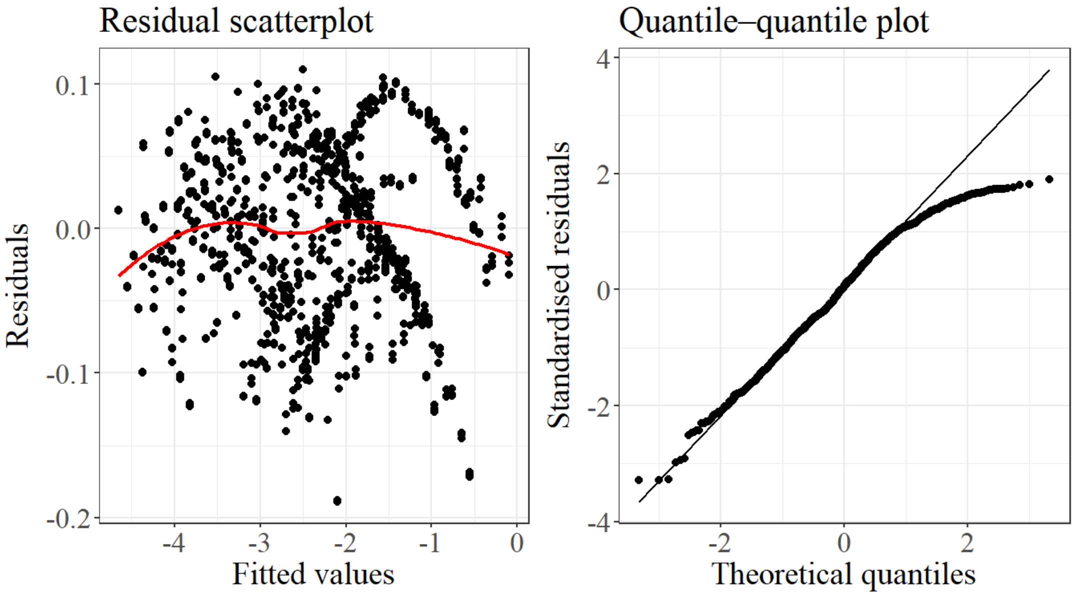


Fig. S5.2 | Diagnostic plots for the $b_0$ model. Residuals versus fitted values and normal Q–Q plot for the linear model applied to log($b_0$). The diagnostics show adequate model behaviour across the central fitted range, with some departure from normality in the upper residual tail.

The slope parameter $k$ is strictly negative and spans orders of magnitude. Modelling ln(−$k$) therefore gives a stable relation across the domain. Its variation with $G$, $r$ and $C_f$ is well described by

$$\ln(-k) = c_0 + c_1 \log_{10}(G) + c_2 \log_{10}(r) + c_3 C_f + c_4 C_f^2 + c_5 \log_{10}(G) C_f + c_6 \log_{10}(r) C_f$$

with $k$ obtained as $-e^{\ln(-k)}$.

This model explains 98.7% of the variance in log(−$k$) in the training dataset, with residual error approximately 0.11.

Table S5.2 Coefficients for the $k$ model (ln(−$k$) scale)

| **Coefficient** | **Estimate** | **StdError** |
|---|---|---|
| Intercept | 3.05724 | 0.18210 |
| $\log_{10}(G)$ | 0.95976 | 0.01584 |
| $\log_{10}(r)$ | -1.05952 | 0.04831 |
| $C_f$ | -0.34032 | 0.30456 |
| $C_f^2$ | -1.88336 | 0.09125 |
| $\log_{10}(G)$:$C_f$ | -0.20556 | 0.02504 |
| $\log_{10}(r)$:$C_f$ | 0.08384 | 0.07639 |
| *Model Residual SE*: 0.1064 | | |
| $R^2$: 0.9869, adj. $R^2$: 0.9868 | | |

Agreement between observed and predicted $k$ remains strong (Fig. S5.3), and residual structure is well behaved (Fig. S5.4).

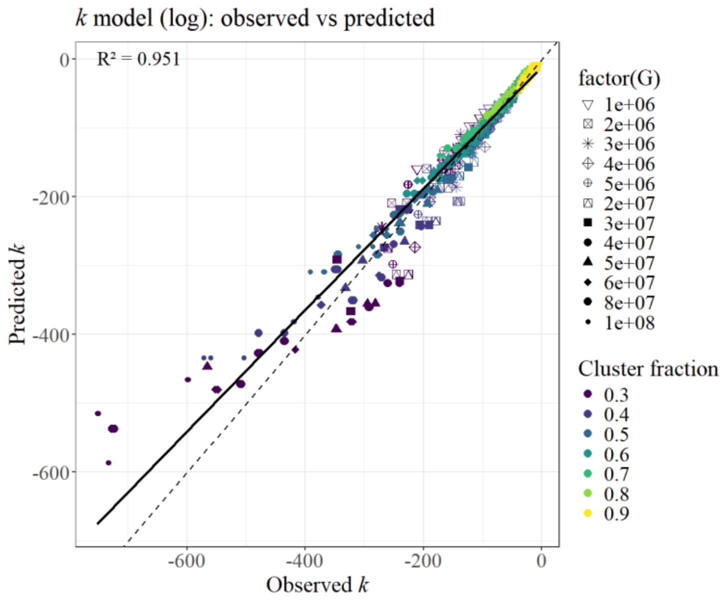


Fig. S5.3 | Observed versus predicted values of $k$. Observed $k$ from calibration run plotted against predicted $k$ obtained from the fitted expression for $\log(-k)$. Colours indicate cluster fraction and shapes indicate genome size. The dashed line denotes the 1:1 relation.

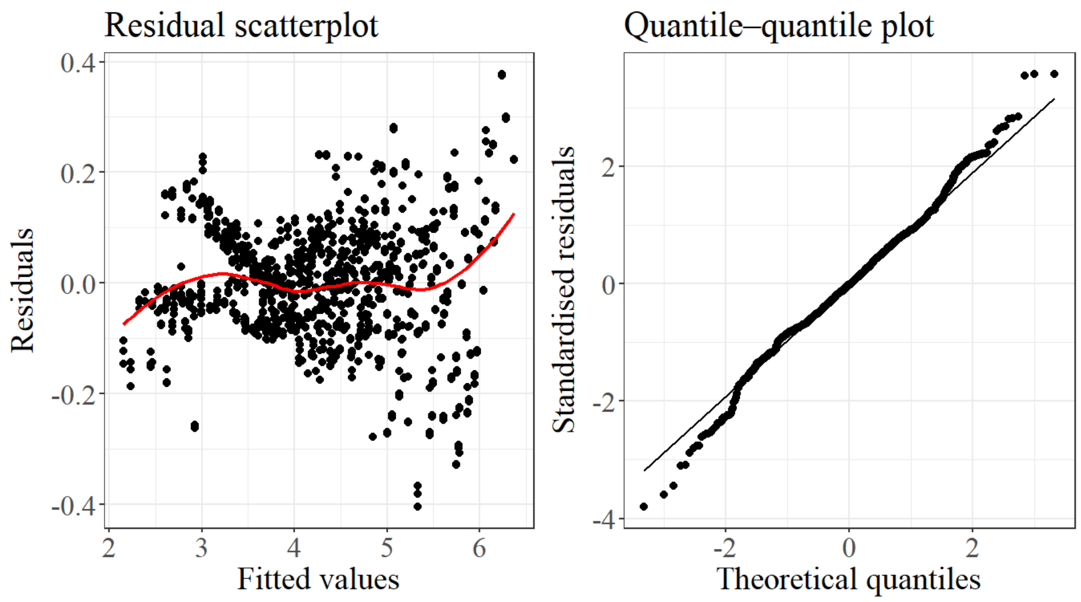


Fig. S5.4 | Diagnostic plots for the k model. Residuals versus fitted values and normal Q–Q plot for the linear model applied to $\log(-k)$. Residual patterns remain broadly consistent with model assumptions across the parameter domain.

The fitted coefficients are reported in Tables S5.1 and S5.2. Both expressions are smooth, monotonic and continuous in all three variables, and therefore suitable for use as reversible components of the logistic curve when computing MAVI values.

Five additional simulation sets (runs 1 to 5), generated independently of the training data, were used to assess robustness. These sets cover the following ranges of parameters:

Table S5.3. Grids of $G$, $r$ and $C_f$ in calibration runs and runs 1 to 5.

| **Run** | **Reps.** | **$G$ (×10^6)** | **$r$ (×10^3)** | **$C_f$** | **Total Runs** |
|---|---|---|---|---|---|
| Cal. 1 | 3 | 1, 2, 3, 4, 5 | 1, 2, 3, 4, 5, 6 | 0.3, 0.4, 0.5, 0.6, 0.7, 0.8, 0.9 | 630 |
| Cal. 2 | 3 | 5, 20, 30, 40, 50, 60, 80, 100 | 3, 4, 5 | 0.3, 0.4, 0.5, 0.6, 0.7, 0.8, 0.9 | 504 |
| 1 | 3 | 0.4, 0.5, 0.6, 0.7, 0.8, 0.9 | 0.6, 0.7, 0.8, 0.9, 1, 2 | 0.3, 0.4, 0.5, 0.6, 0.7, 0.8 | 648 |
| 2 | 3 | 2, 4, 6, 8, 10 | 2, 5, 8, 10, 15 | 0.3, 0.4, 0.5, 0.6, 0.7, 0.8, 0.9 | 525 |
| 3 | 3 | 0.5, 1, 2, 3, 4, 5, 6 | 1, 2, 3, 4, 5, 6, 7 | 0.3, 0.4, 0.5, 0.6, 0.7, 0.8, 0.9 | 1029 |
| 4 | 3 | 1, 2, 3, 4, 5, 6, 8 | 1, 2, 3, 4, 5, 6, 7 | 0.3, 0.4, 0.5, 0.6, 0.7, 0.8, 0.9 | 1029 |
| 5 | 3 | 20, 30, 40, 50, 60, 80, 100 | 3, 4, 5, 6, 7, 10, 15 | 0.3, 0.4, 0.5, 0.6, 0.7, 0.8, 0.9 | 1029 |

Prediction accuracy remained high across all runs (Table S5.4). For $b_0$, $R^2$ values ranged from 0.97 to 0.99, with very small biases. For $k$, $R^2$ ranged from approximately 0.9 to 0.96, reflecting both the greater stochastic variability of $k$ and the narrower scale on which $k$ acts, but with low mean bias in all cases. Cross-run performance is summarised in Fig. S5.5.

Table S5.4. Predictive accuracy in independent validation runs.

| **Run** | **n** | **$R^2$ $b_0$** | **RMSE $b_0$** | **Bias $b_0$** | **$R^2$ $k$** | **RMSE $k$** | **Bias $k$** |
|---|---|---|---|---|---|---|---|
| 1 | 648 | 0.965 | 0.0144 | 0.00906 | 0.897 | 16.9 | 8.74 |
| 2 | 525 | 0.971 | 0.0301 | 0.0117 | 0.944 | 12.4 | 0.181 |
| 3 | 1029 | 0.981 | 0.0277 | 0.00546 | 0.963 | 10.4 | 0.605 |
| 4 | 1029 | 0.991 | 0.0158 | 0.00150 | 0.961 | 11.9 | -0.693 |
| 5 | 1029 | 0.991 | 0.00759 | 0.000135 | 0.931 | 32.4 | -1.83 |

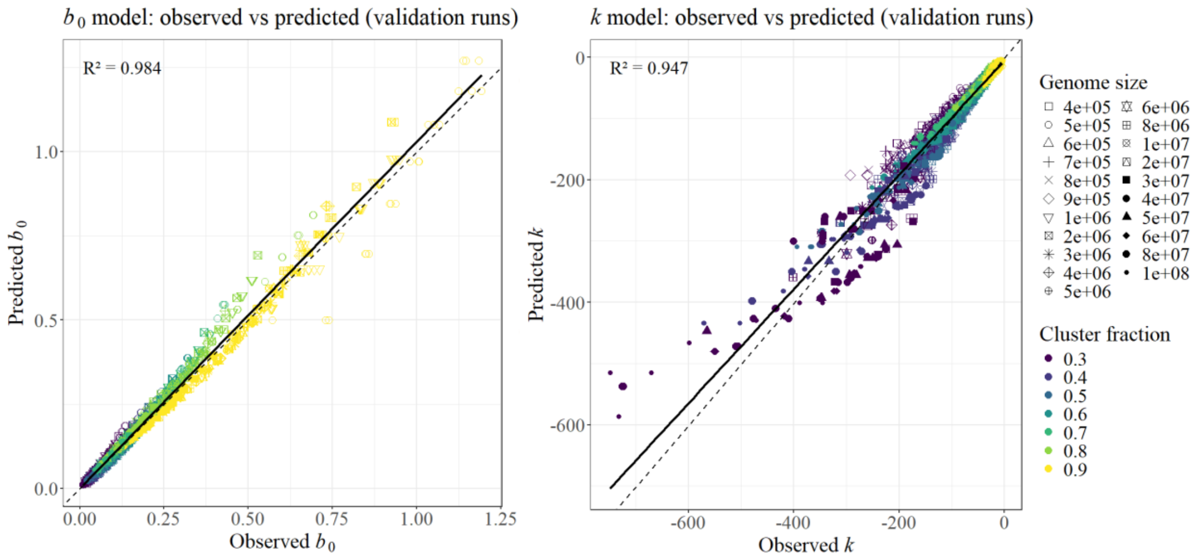


Fig. S5.5 | Cross-run predictive accuracy of the $b_0$ and $k$ models.

These results show that the two parametric forms $b_0(G, r, C_f)$ and $k(G, r, C_f)$ provide accurate and computationally efficient approximations to the logistic parameters across the full simulation range. The equations reproduce the simulation outputs with high fidelity and generalise well to independent datasets, allowing the logistic model to be applied directly when evaluating genomic presence using the MAVI framework.

**Relevance to MAVI**

These equations allow the logistic curve to be reconstructed directly from $G$, $r$ and $C_f$, which makes the statistical test fully reversible and therefore usable within the MAVI framework. Their importance is practical: $b_0(G, r, C_f)$ and $k(G, r, C_f)$ describe how rapidly the $\beta_0$-controlled compatibility boundary changes and where its transition occurs, so monotonic and consistent behaviour guarantees that the mapping between breadth and the compatibility boundary is stable across datasets. The exact algebraic form is unimportant. What matters is that the functions reproduce the simulated behaviour with high accuracy, allowing MAVI to compute support-fraction bounds without rerunning the full clustered simulations.

### Supplementary Note 6 – Minimax identifiability and the Metagenomic Alignment Validation Index

The Generator Identifiability Envelope (GIE) introduced by Márquez and Silva-Toro [2] provides the admissible minimax support-fraction bound for stochastic generators subject to unobservable support restriction. For a declared dispersion observable and mimicry tolerance $\beta_0$, the GIE reports the supremal support-fraction bound that can be uniformly ruled out as insufficient to reproduce the observed full-support compatibility under the worst-case admissible restricted-support alternative. It therefore provides a conservative lower bound on the fraction of the generator domain required by the observed evidence.

Under the metagenomic specialisation, the reference domain Ω is the unit-normalised candidate genome, and the Boolean germ–grain process corresponds to the stochastic survival and placement of genomic fragments. The stabilising dispersion functional $D_s$ is instantiated as the alignment-block dispersion statistic $D$, calculated after merging reads separated by less than the declared genomic scale $s$.

The full-support null law of $D$ induces the compatibility coordinate $Y = F_0(D)$, where $F_0$ is the cumulative distribution function of dispersion under random full-genome fragmentation.

Operationally, this coordinate is represented by $Y_{\text{obs}} = \Phi(Z_{\text{obs}})$, where $Z_{\text{obs}}$ is obtained by standardising the observed dispersion against the corresponding Monte Carlo null distribution.

For $c \in (0,1]$, let

$$\mathcal{A}_c = \left\{ A \subseteq \Omega : A \text{ is admissible and } \frac{|A|}{|\Omega|} \le c \right\}$$

denote the class of admissible restricted genomic supports occupying at most fraction *c* of the candidate genome. Suppressing the fixed values of *G*, *r*, and *s*, the worst-case probability that such a support reproduces a compatibility value above *t* at breadth *b* is

$$\beta^*(c,t \mid b) = \sup_{A \in \mathcal{A}_c} \Pr_A \{ Y_b > t \}.$$

For a declared tolerance $\beta_0$, the corresponding restricted-support compatibility boundary is

$$T(b \mid c) = \inf \{ t \in [0,1] : \beta^*(c,t \mid b) \le \beta_0 \}.$$

Thus, $T(b \mid c)$ is the level of full-support compatibility that generators operating on support fractions no greater than *c* can exceed with probability at most $\beta_0$, uniformly over the admissible restricted-support class.

For an observed candidate with breadth $b_{\text{obs}}$ and compatibility coordinate $Y_{\text{obs}} = \Phi(Z_{\text{obs}})$, the genomic support-fraction envelope is

$$C_f^* = \sup \{ c \in (0,1] : \Phi(Z_{\text{obs}}) > T(b_{\text{obs}} \mid c) \}.$$

If $\Phi(Z_{\text{obs}}) > T(b_{\text{obs}} \mid c)$, then restricted supports occupying at most fraction *c* reproduce the observed compatibility only within the declared worst-case mimicry tolerance. Such support fractions are therefore ruled out as insufficient explanations of the observed dispersion evidence. Taking the supremum identifies the boundary of this ruled-out region.

In practice, the theoretical boundary $T(b \mid c)$ is replaced by the fitted empirical boundary $\hat{T}(b \mid c)$ developed in Supplementary Notes 1–5. The Metagenomic Alignment Validation Index is therefore defined as

$$\mathrm{MAVI} = \sup\left\{c \in (0,1] : \Phi(Z_{\mathrm{obs}}) > \hat{T}(b_{\mathrm{obs}} \mid c)\right\}.$$

MAVI is consequently the empirical genomic specialisation of the Generator Identifiability Envelope. It reports the supremal genomic support-fraction bound that can be ruled out as too small to explain the observed breadth–dispersion evidence under the declared worst-case mimicry tolerance. Equivalently, it provides a conservative lower bound on the fraction of the candidate genome required for a restricted-support explanation to remain viable.

**Relevance to MAVI**
Thus, MAVI is the one-dimensional biological specialisation of the Generator Identifiability Envelope.

**Supplementary Note 7 – Hierarchical generator-level dispersion laws and workflow entropy**

Although the operational development of MAVI foregrounds the Generator Identifiability Envelope (GIE), the GIE is itself downstream of the generator-level dispersion law established by Márquez and Silva-Toro [2]. For a stochastic generator $U$, a realised recovery $X_U$ gives the stabilising dispersion observable $D_s(X_U)$, and repeated interrogation induces the generator-level dispersion law

$$\mathcal{L}_s(U) = \mathrm{Law}\left(D_s(X_U)\right).$$

This law, rather than any individual recovery, is the generator-level object from which both dispersion entropy and support-fraction inversion are constructed[1,2]. This law also admits a natural hierarchical construction through standard probability and information theory.

A population-level random mechanism may generate observations through a family of conditional laws indexed by a context variable Λ, with the resulting marginal law obtained by averaging those conditional laws over the distribution of Λ. Once a common observable $Y$ is defined, any finite representation $J = q_B(Y)$ obeys the classical Shannon decomposition

$$H(Y) = H(J \mid \Lambda) + I(J;\Lambda),$$

where $H(J \mid \Lambda)$ measures the remaining variability within conditional contexts and $I(J; \Lambda)$ measures the information carried by $J$ about which context generated it, following the classical Shannon decomposition[3]. The use of a workflow-level law therefore requires no new information-theoretic principle; the required hierarchical and entropy structure is already classical.

Under the metagenomic specialisation, genomic coordinate replaces Euclidean space, surviving aligned fragments replace recovered Boolean grains, and $D_s$ is instantiated by the alignment-block dispersion statistic $D$.

For an individual candidate, let λ denote its recovery context, including the candidate reference genome and the quantities that determine its conditional recovery law, such as genome length $G$, mapped-read count $x$ and empirical read-length distribution $r$. Conditional on this context, the candidate has a generator $U_\lambda$ and corresponding dispersion law

$$U_\lambda \to X_\lambda \to D_s(X_\lambda) \to \mathcal{L}_{s,\lambda}, \qquad \mathcal{L}_{s,\lambda} = \mathrm{Law}\big(D_s(X_\lambda) \mid \Lambda = \lambda\big).$$

The candidate-level GIE construction developed in Supplementary Notes 1-6 acts on this surviving dispersion structure to infer how much genomic support is required by the observed evidence. MAVI is therefore the inverse branch of a construction whose primary observable object is the candidate-level dispersion law.

#### 7.1 Hierarchical workflow generator

A metagenomic workflow produces many candidate recoveries across samples. These candidate generators form the conditional components of a higher-level workflow generator.

Let

$$\Lambda \sim \Pi_{\mathrm{wf}}$$

denote the candidate context generated within a declared biological and analytical workflow. Conditional on $\Lambda = \lambda$, recovery follows $U_\lambda$. The workflow generator can therefore be represented hierarchically as

$$U_{\mathrm{wf}} = \big(\Pi_{\mathrm{wf}}, \{U_\lambda\}_\lambda\big).$$

A realisation of the workflow-level generator follows the hierarchy

$$U_{\mathrm{wf}} \to \Lambda \to U_\Lambda \to X_\Lambda \to D_s(X_\Lambda).$$

Raw dispersion values are candidate-specific because their full-support reference laws depend on the recovery context, including $G$, $x$ and $r$. Direct pooling of $D_s$ across candidates would therefore mix observables defined on different reference scales. MAVI resolves this by mapping each candidate dispersion state to its candidate-specific null-standardised compatibility coordinate

$$Y_\lambda = \psi_\lambda\big(D_s(X_\lambda)\big) = \Phi(Z_\lambda),$$

where

$$Z_\lambda = \frac{D_s(X_\lambda) - \mu_{0,\lambda}}{\sigma_{0,\lambda}}.$$

For $\sigma_{0,\lambda} > 0$, $\psi_\lambda$ is strictly monotone in $D_s$ and therefore preserves the ordering of the candidate-level dispersion states while placing them on the common interval [0,1]. At the level of probability laws,

$$\mathcal{L}_\lambda^Y = (\psi_\lambda)_\# \mathcal{L}_{s,\lambda},$$

where $(\psi_\lambda)_\#$ denotes the pushforward of the candidate-level dispersion law through its compatibility transformation.

The workflow consequently induces the common observable law

$$\mathcal{L}_{\mathrm{wf}}^Y = \mathrm{Law}(Y) = \int (\psi_\lambda)_\# \mathcal{L}_{s,\lambda}\, \Pi_{\mathrm{wf}}(d\lambda),$$

across candidate recoveries generated by the workflow. The cohort distribution of $\Phi(Z)$ is therefore the marginal observable law of a hierarchical generator whose candidate-specific dispersion laws are mapped to a common compatibility coordinate before being combined according to the workflow distribution $\Pi_{\mathrm{wf}}$.

The structure may be summarised as

$$U_{\mathrm{wf}} \to \{U_\lambda \to X_\lambda \to D_s(X_\lambda) \to \mathcal{L}_{s,\lambda}\}_{\lambda\sim\Pi_{\mathrm{wf}}} \to Y \to \mathcal{L}_{\mathrm{wf}}^Y.$$

The candidate-level GIE and the workflow-level entropy developed below are therefore two interrogations of the same hierarchy of generator-level dispersion laws.

#### 7.2 Empirical cohort entropy

The cohort analysis estimates the workflow-level observable law from candidate recoveries retained for the entropy analysis. Let $R = 1$ denote that a candidate passes the operational $\alpha$-compatibility gate and possesses a data-derived compatibility coordinate.

The relevant workflow law is therefore the retained law

$$\mathcal{L}_{\mathrm{wf}}^{Y,\mathrm{ret}} = \mathrm{Law}(Y \mid R = 1); \qquad \mathcal{L}_{\mathrm{wf}}^{J,\mathrm{ret}} = (q_B)_{\#}\mathcal{L}_{\mathrm{wf}}^{Y,\mathrm{ret}}, \qquad J = q_B(Y).$$

For the $n$ retained candidates in a cohort, the interval $[\alpha,1]$ is partitioned into $B = 25$ equal-width bins. Let

$$\hat{p}_j = \frac{1}{n}\sum_{i=1}^{n} \mathbf{1}\{q_B(Y_i) = j\}, \qquad j = 1,\dots,B$$

The empirical workflow entropy is

$$\hat{H}_{\mathrm{wf}} = -\sum_{j=1}^{B} \hat{p}_j \log \hat{p}_j$$

and the normalised entropy reported in the manuscript is

$$H_{\mathrm{norm}} = \frac{\hat{H}_{\mathrm{wf}}}{\log B} \qquad \in [0,1]$$

At this declared resolution, $H_{\mathrm{norm}} = 0$ exactly when all retained candidate recoveries occupy the same compatibility bin. Increasing values indicate that the workflow-level observable law is distributed across a larger and/or more evenly occupied set of compatibility states. A uniform distribution over all $B$ states gives $H_{\mathrm{norm}} = 1$.

Because $H(J) = H(J \mid \Lambda) + I(J; \Lambda)$ with both terms non-negative, entropy collapse implies loss of observable variation both within candidate contexts and among the contexts represented by the workflow.

The empirical quantity is therefore the entropy of the candidate-weighted observable law emitted by the workflow within the analysed cohort. It is not a measure of taxonomic diversity, nor does its construction require all candidates to have identical genomes, read counts or read-length distributions. Those differences belong to the context variable $\Lambda$, while the candidate-specific transformation $Y = \Phi(Z)$ places the surviving dispersion evidence on the common coordinate used to construct the workflow-level law.

The relation between the candidate-level and workflow-level branches can be summarised as

$$\begin{aligned} &U_\lambda \to X_\lambda \to D_s(X_\lambda) \to \mathcal{L}_{s,\lambda}, \\ &\mathcal{L}_{s,\lambda} \to \text{GIE inversion} \to \text{MAVI}_\lambda, \\ &\{\mathcal{L}_{s,\lambda}\}_{\lambda\sim\Pi_{\text{wf}}} \xrightarrow{\psi_\lambda} \mathcal{L}_{\text{wf}}^{Y,\text{ret}} \xrightarrow{q_B} \mathcal{L}_{\text{wf}}^{J,\text{ret}} \to H_{\text{norm}} \end{aligned}$$

Thus, MAVI and the cohort entropy both descend from the generator-level dispersion law established under partial observation. GIE inversion interrogates the candidate-level law to obtain a conservative genomic support bound, whereas workflow entropy interrogates the hierarchical law induced across candidate recoveries to determine whether observable dispersion variation survives at cohort level.

**Relevance to MAVI**

The generator-level dispersion law is the common structural foundation of the candidate- and workflow-level analyses. Its candidate-level inverse structure yields MAVI, while its hierarchical workflow composition yields the cohort entropy used to detect collapse of observable dispersion variation.

### Supplementary Note 8 – Read-support inclusion and minimum validated community explanations

Let $V = \{1, \ldots ,p\}$ denote the set of MAVI-retained taxa for a sample. For each taxon $i \in V$, let

$$R_i \subseteq R$$

be the set of unique observed read identifiers contributing to its mapped evidence, where $R$ is the read universe entering the community analysis. The complete read evidence represented by the retained candidate set is

$$R_V = \bigcup_{i\in V} R_i \,.$$

For the reporting threshold $\tau$, let

$$V_\tau = \{i \in V : m_i \geq \tau\}$$

denote the subset of retained candidates at or above that threshold, with corresponding read evidence

$$R_{V_\tau} = \bigcup_{i \in V_\tau} R_i .$$

#### 8.1 Read-support equivalence and inclusion

The family

$$\mathcal{R}_V = \{R_i : i \in V\}$$

inherits the ordinary inclusion relation of sets. Two taxa have identical read support when

$$i \sim j \quad \Leftrightarrow \quad R_i = R_j .$$

Equality therefore partitions the candidate set into read-support equivalence classes. On these classes, inclusion induces the partial order

$$[i] \preceq [j] \quad \Leftrightarrow \quad R_i \subseteq R_j .$$

Thus, if $R_i \subset R_j$, every read represented by $i$ is already represented by $j$, whereas $R_i = R_j$ means that the two taxa cannot be distinguished from read membership alone.

The corresponding MAVI value $m_i$ is retained as a secondary candidate-level coordinate. It does not define the inclusion order: candidates with identical read sets may have different MAVI values because those reads can occupy different genomic positions relative to different reference genomes.

#### 8.2 Counterfactual necessity

For each candidate $i$, define its uniquely contributed read set

$$U_i = R_i \setminus \bigcup_{j \in V \setminus \{i\}} R_j .$$

If $U_i \neq \varnothing$, then removal of candidate $i$ leaves at least one read in $R_V$ unexplained. Consequently, $i$ must occur in every exact explanation of the complete validated read set.

This provides a sufficient counterfactual condition for necessity: a candidate with uniquely contributed read evidence cannot be removed from any complete explanation. However, a candidate may also be necessary in every minimum-cardinality explanation even when $U_i \neq \varnothing$, if removing it requires a larger combination of alternative taxa. Necessity in the minimum community is therefore defined below from membership across minimum solutions.

#### 8.3 Minimum validated community explanation

For any subset $C \subseteq V$, define its explained-read set as

$$R(C) = \bigcup_{i \in C} R_i \,.$$

The minimum community structure is evaluated first within $V_\tau$. The family of minimum-cardinality explanations of the read evidence represented at or above the threshold is An exact minimum validated community explanation is any solution of

$$\mathcal{C}_\tau^* = \arg\min_{C \subseteq V_\tau} |C| \quad \text{subject to} \quad R(C) = R_{V_\tau}$$

The corresponding minimum validated community is then obtained after admitting the complete retained candidate set $V$:

$$\boxed{\mathcal{C}^* = \arg\min_{C \subseteq V} |C| \quad \text{subject to} \quad R(C) = R_V .}$$

Neither solution need be unique. Thus $\mathcal{C}^*$ and $\mathcal{C}_\tau^*$ denote the families of minimum-cardinality taxonomic configurations capable of explaining $R_{V_\tau}$ and $R_V$, respectively.

Taxa at or above the reporting threshold that occur in every minimum explanation at both levels form the necessary set

$$N^* = V_\tau \cap \left( \bigcap_{C \in \mathcal{C}_\tau^*} C \right) \cap \left( \bigcap_{C \in \mathcal{C}^*} C \right).$$

These taxa remain necessary even after lower-$m_i$ retained candidates are admitted as alternative explanations.

A taxon $i \in V_\tau$ is redundant when

$$i \notin \bigcup_{C \in \mathcal{C}_\tau^*} C \,,$$

because it is absent from every minimum explanation even before candidates below the reporting threshold are admitted.

The remaining taxa at or above the threshold form the competing set

$$Q^* = \left( V_\tau \cap \bigcup_{C \in \mathcal{C}_\tau^*} C \right) \setminus N^* .$$

These taxa participate in a minimum explanation of $R_{V_\tau}$, but their taxonomic identity is not uniquely required after all retained alternatives are considered. This includes both alternative minimum explanations within $V_\tau$ and taxa that become unnecessary only after candidates below the threshold are admitted.

The corresponding MAVI value $m_i$ provides an independent measure of the genomic support associated with each candidate. Necessary candidates below the reporting threshold are therefore defined as

$$N_{\text{res}} = \left\{ i \in \bigcap_{C \in C^*} C : m_i < \tau \right\}.$$

These residual necessary candidates are required to account for the complete observed read evidence, but their low MAVI values provide insufficient support for confident species-level interpretation. Their contribution can therefore be quantified directly from the read structure.

The read evidence that can only be represented after candidates below the reporting threshold are admitted is

$$R_{\text{res}} = R_V \setminus R_{V_\tau}.$$

The corresponding residual read fraction, relative to the read universe entering the community analysis, is

$$\rho_{\text{res}} = \frac{|R_{\text{res}}|}{|R|}.$$

$\rho_{\text{res}}$ therefore quantifies the fraction of the aligned read evidence that cannot be represented by candidates in $V_\tau$ and requires the lower-$m_i$ retained layer.

Within this residual evidence, the reads uniquely contributed by residual necessary taxa are

$$R_{\text{priv,res}} = \bigcup_{i \in N_{\text{res}}} U_i,$$

with fraction

$$\rho_{\text{priv,res}} = \frac{|R_{\text{priv,res}}|}{|R|}.$$

The remaining residual evidence

$$\rho_{\mathrm{res}} - \rho_{\mathrm{priv,res}}$$

is shared among multiple retained candidates and therefore cannot be uniquely assigned from read membership alone.

Residual necessary candidates are retained explicitly in the minimum explanation rather than discarded, allowing their identity and contribution to be inspected directly. MAVI provides the genomic-support coordinate on the read-support structure.

### References


1. Márquez, D. A. & Silva-Toro, F. Generator-level dispersion laws for Boolean random closed sets under partial observation. (2026). https://doi.org/10.5281/zenodo.22884521
2. Márquez, D. A. & Silva-Toro, F. Generator Identifiability Envelope: Minimax support-fraction inference for Boolean random closed sets under hidden support restriction. (2026). https://doi.org/10.5281/zenodo.22884693
3. Shannon, C. E. A mathematical theory of communication. *The Bell System Technical Journal* **27**, 379-423 (1948). https://doi.org/10.1002/j.1538-7305.1948.tb01338.x
